# BA.1 and BA.2 sub-lineages of Omicron variant have comparable replication kinetics and susceptibility to neutralization by antibodies

**DOI:** 10.1101/2022.01.28.22269990

**Authors:** Janmejay Singh, Aleksha Panwar, Anbalagan Anantharaj, Chitra Rani, Monika Bhardwaj, Parveen Kumar, Kamal Pargai, Partha Chattopadhyay, Priti Devi, Ranjeet Maurya, Pallavi Mishra, Anil Kumar Pandey, Rajesh Pandey, Guruprasad R. Medigeshi

## Abstract

The Omicron variant of SARS-CoV-2 is capable of infecting unvaccinated, vaccinated and previously-infected individuals due to its ability to evade neutralization by antibodies. With three sub-lineages of Omicron emerging in the last four months, there is inadequate information on the quantitative antibody response generated upon natural infection with Omicron variant and whether these antibodies offer cross-protection against other sub-lineages of Omicron variant. In this study, we characterized the growth kinetics of Kappa, Delta and Omicron variants of SARS-CoV-2 in Calu-3 cells. Relatively higher amounts infectious virus titers, cytopathic effect and disruption of epithelial barrier functions was observed with Delta variant whereas infection with Omicron variant led to a more robust induction of interferon pathway, lower level of virus replication and mild effect on epithelial barrier. The replication kinetics of BA.1 and BA.2 sub-lineages of the Omicron variant were comparable in cell culture and natural Omicron infection in a subset of individuals led to a significant increase in binding and neutralizing antibodies to both BA.1 and BA.2 sub-lineages but these levels were lower than that produced against the Delta variant. Finally, we show that Cu^2+^, Zn^2+^ and Fe^2+^ salts inhibited *in vitro* RdRp activity but only Cu^2+^ and Fe^2+^ inhibited both the Delta and Omicron variants in cell culture. Thus, our results suggest that high levels of interferons induced upon infection with Omicron variant may counter virus replication and spread. Waning neutralizing antibody titers rendered subjects susceptible to infection by Omicron variant and natural Omicron infection elicits neutralizing antibodies that can cross-react with other sub-lineages of Omicron and other variants of concern.

## INTRODUCTION

Close to 17 million cases of COVID-19 was reported from India during the second wave of SARS-CoV-2 from March 2021 to June 2021 overwhelming the public health infrastructure resulting in close to 40% of the >500,000 deaths that has occurred in this pandemic. The surge was due to novel SARS-CoV-2 variant from the lineage B.1.617.2 (Delta variant) which was shown to be more virulent with shorter incubation periods and ability to cause severe disease(1-4). Many reports have shown that the neutralizing antibodies from prior infection or that elicited by some of the licensed COVID-19 vaccines show a decrease in the efficiency to neutralize the Delta variant thereby compromising vaccine efficacy against this variant(5-7). Delta variant was the major circulating SARS-CoV-2 variant until the emergence of B.1.1.529 (Omicron) variant in November 2021 which has now replaced the Delta variant in most of the countries. The ongoing third wave of SARS-CoV-2 infections, which is on the decline and attributed to the Omicron variant of SARS-CoV-2, has resulted in milder symptoms and lower hospitalizations. In addition to the less virulent Omicron variant, increased vaccination coverage and hybrid immunity may have contributed to reduced impact of third wave in India. Three sub-lineages of Omicron (BA.1, BA.2 and BA.3) have emerged and are currently circulating in India and other parts of the world(8). The success of Omicron variant and its sub-lineages in driving the current phase of the pandemic has been attributed to a large number of mutations leading to escape from neutralizing antibodies generated by prior infection or vaccination(9-12). Recent reports suggest that the antibody response between the sub-lineages may not be significantly different and exposure to one of the sub-lineages offers cross-protection against other members of Omicron sub-lineage and also against past variants(13, 14).

In addition to evading humoral immunity, data from animal models suggest that the success of SARS-CoV-2 variants of concern (VoCs) in driving new waves of infection can be attributed to multiple factors such as increased virulence, enhanced transmission, altered pathogenicity and replication fitness(15-20). Therefore, efforts to characterize and understand the replication kinetics of circulating VoCs would provide information on virulence mechanisms, immune evasion properties and augment our ability to develop therapeutic strategies. In this study, we compared the replication kinetics of some of the SARS-CoV-2 VoCs namely, the Kappa and Delta variants and BA.1 and BA.2 sub-lineages of Omicron variant in human lung epithelial adenocarcinoma (Calu-3) cells grown on transwells and also in air-liquid interface models. We monitored the interferon response and its downstream effectors in infected cells and tested the effect of divalent cations on virus infection and RNA-dependent RNA polymerase (RdRp) activity. We show that both binding and neutralizing antibodies wane by >50% in six months in a cohort of participants with hybrid immunity. Natural exposure to the Omicron variant led to a significant increase in binding and neutralizing antibodies to BA.1 and BA.2 sub-lineages of Omicron. The levels of neutralizing antibodies were higher against the Delta variant as compared to the Omicron sub-lineages. These data suggest that Omicron infection elicits neutralizing antibodies that can cross-react with other sub-lineages of Omicron and other VoCs in people with hybrid immunity. Our results provide clues to the relative replication fitness of these VoCs and their ability to induce interferon responses and susceptibility to divalent cations and may help to develop novel strategies to counter viral replication.

## RESULTS

### Peak COVID-19 positivity in second wave coincides with high viral load

A total of 117,434 NP/OP swab samples were tested for COVID-19 by RT-PCR at the bioassay laboratory of the institute from April 2020 to July 2021. Peak positivity in the year 2020 was observed in the months of June-July with 15% positivity during these months (Supplementary Fig. S1A(i)). Previous reports have identified B.6 lineage as one of the predominant lineages during this period circulating in India (21, 22). We have isolated few clinical isolates from B.6 lineage as reported earlier and plaque-purified one of the isolates which was used in this study as a representative of ancestral virus isolate (23). The number of positive cases was around 10% of the total samples tested until November 2020 and dropped drastically to around 0.1% of the total tested samples in the month of February 2021 (Supplementary Fig. S1A(ii)). Although only 8 of the 8012 tested samples were RT-PCR positive in February 2021, 4 of these 8 samples had RT-PCR cycle threshold (Ct) values <20 and 2 with Ct values between 20-25. RT-PCR positivity jumped to 1.1% (85 out of 7558) in March with over 50% of the positive samples having Ct values of <25 clearly indicating a large number of infections with a high viral load during February and March 2021 (Supplementary Fig. S1B). The following months of April and May saw a sudden spike in the peak positivity rate which increased to 27.7% and 24.5% which is now termed as the “second wave” and the SARS-CoV-2 variants from B.1.617.1/2 (Kappa/Delta) lineages have been attributed to this sudden spike which overwhelmed the healthcare infrastructure leading to large scale mortality (24). 40% of the NP/OP samples tested in April 2021 had Ct values of less than 25 which reduced to 28% of the total samples in May indicating a high viral burden in patients during this period resulting in increased transmission of the virus among the contacts.

### Disruption of cellular junctions by SARS-CoV-2 Delta and Kappa variants

We were able to isolate the B.1.617.1 (Kappa) and B.1.617.2 (Delta) variants from clinical samples and got the virus stocks verified by whole genome sequencing. We measured the growth kinetics of these two variants in comparison with the B.6 lineage virus. Calu-3 cells were infected with 0.01 MOI of SARS-CoV-2 from lineages B.6, B.1.617.1 and B.1.617.2 respectively. We estimated viral titers in the supernatants at 24 and 48 h pi by plaque assay. Total RNA was isolated from cells at each time point for measuring copy numbers of N gene by quantitative RT-PCR. We observed a significantly higher viral titers with B.1.617.2 variant relative to B.6 lineage virus at 24 h pi. The virus titers at 48 h pi was reduced in both the Kappa and the Delta variant samples most probably due to the increased cytopathic effect at this time point (Fig. 1A). However, no difference in N gene RNA copy numbers were detected between the three isolates by RT-PCR at either of the time-points (Fig. 1B). As SARS-CoV-2 was shown to disrupt tight junctions in Calu-3 and human airway epithelial cells (23, 25), we further assessed the effect of infection of B.6 and B.1.617.1 and B.1.617.2 variants on cellular junctions of polarized Calu-3 cells. Cells were grown on transwell culture inserts for 21 days to allow for polarization into apical and basolateral surface and formation of permeability barrier as measured by the trans-epithelial electrical resistance (TEER). Polarized Calu-3 cells were infected with 0.3 MOI of each of the three virus isolates and the TEER was monitored for 48 hours. Cells that were either mock-infected or infected with B.6 lineage virus showed no significant change in TEER up to 48 h (Fig. 1C). Epithelial barrier function was disrupted by 48 h in cells infected with either the Kappa (B.1.617.1) or the Delta (B.1.617.2) variants. However, a significant decline in the TEER values was observed for the Delta variant (Fig.1C). Surprisingly, the virus titers in either the apical or basolateral chambers were not significantly different between the three isolates (Fig. 1D) at 48 h pi suggesting that the increasing cytopathogenicity by the Delta variant may be responsible for significantly higher disruption of lung epithelial barrier functions. These findings were further corroborated by confocal microscopy where transwell culture inserts were fixed at 48 h pi and stained with antibodies against SARS-CoV-2 nucleocapsid (N) protein, occludin (marker for tight junction) and β-catenin (marker for adherens junction) (Fig. 2). The staining pattern of both occludin and β-catenin was comparable between mock- and B.6-infected cells. However, the same was perturbed in both the Kappa and the Delta variant infected cells and disruption of occludin staining appeared to be more drastic in cells infected with the Delta variant compared to the Kappa variant (Fig. 2). Our results suggest that the Delta variant infection has relatively more deleterious effects on epithelial cell functions most probably due to increased fusogenic functions leading to syncytia formation and cell death(19).

**Figure 1:**
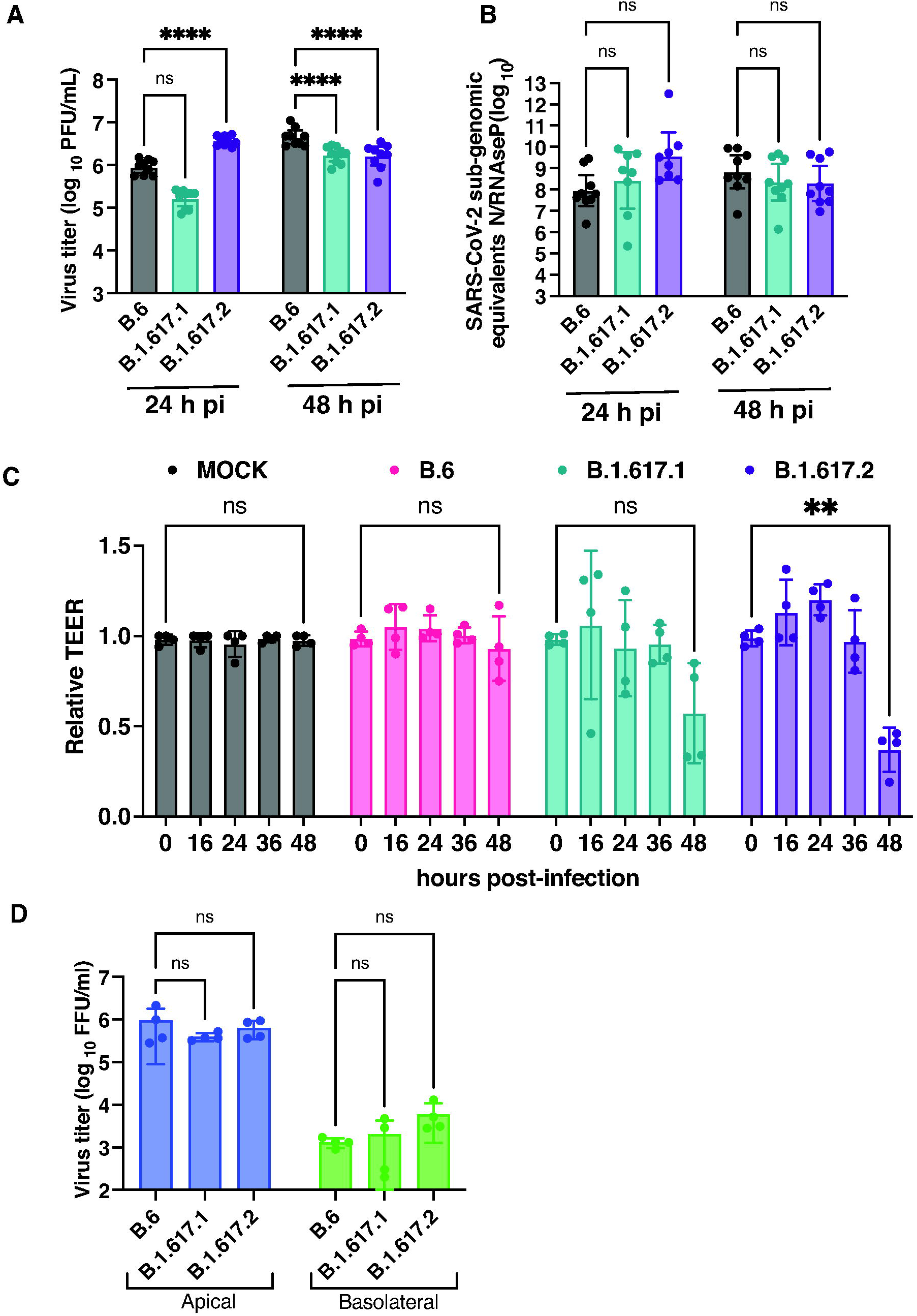
Growth curve analysis of B6, B.1.617.1, and B.1.617.2 variants in Calu-3 cells. Calu-3 cells were infected with 0.01 MOI of SARS-CoV-2 virus B.6, B.1.617.1 (Kappa), and B.1.617.2 (Delta) variants. (A) Virus titer in the culture supernatant collected at indicated time-points were estimated by plaque assay. Error bars represent geometric mean with 95% CI. (B) Sub-genomic RNA of N gene was estimated by qRT-PCR. *RNAse P* was used as internal control for normalization. Error bars represent geometric mean with 95% CI. (C) Calu-3 cells grown on transwell inserts were infected with above variants at 0.3 MOI and TEER was monitored at indicated time points. Values (Mean ± SD) are represented as relative to time zero before infection and statistical significance was determined by comparing the zero hour and 48 h readings by two-way ANOVA with Dunnett’s multiple comparisons test. (D) Viral titers were measured in supernatants at 48 h pi by focus-forming units. Data are from two or three independent experiments (Mean ± SD). ns: non-significant; ** P<0.01; **** P<0.0001.

**Figure 2:**
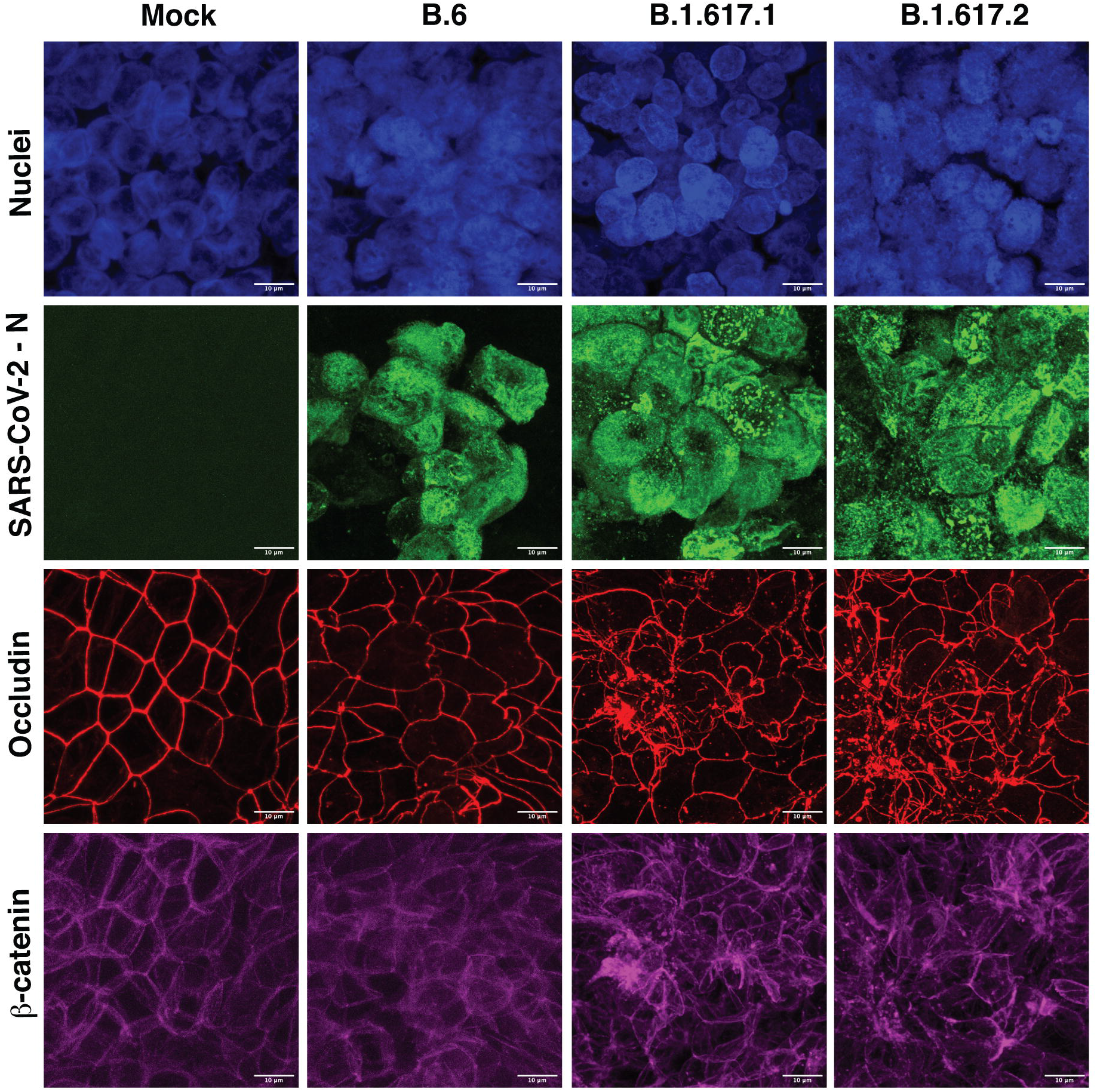
Kappa and Delta variants of SARS-CoV-2 disrupt epithelial junctions. Calu-3 cells were grown on transwell inserts and infected with indicated variants of SARS-CoV-2 at 0.3 MOI. At 48 h pi, cells were fixed and stained with occludin and β-catenin antibodies. Nucleocapsid antibody was used to visualize the SARS-CoV-2 infection. Appropriate Alexa Fluor dye-conjugated secondary antibodies were used for visualization. Nuclei were stained with DAPI. Images were captured at 100X magnification. Images were analyzed using cellSens software and Z-projection at maximum intensity images are shown in the figure. Scale bar is 10 µM.

### Infection with Omicron variant generates lower levels of infectious virus

Delta variant was reported to have higher transmissibility and pathogenicity in humans and animal models(19, 26-28) whereas Omicron variant showed attenuated growth(17, 29). We performed growth curve analysis of SARS-CoV-2 viruses from the lineages B.6 (isolate from 2020), B.1.617.2 (Delta) and B.1.1.529 (Omicron) in Calu-3 cells by infecting cells with 0.3 MOI and estimating viral genome equivalents (N gene) by qRT-PCR from infected cells and virus titers from culture supernatants by plaque assays at indicated time-points (Fig. 3). We found that viral RNA replication peaked by 24 h pi for both the B.6 and the Delta variants and both these isolates had comparable viral N-gene levels by qRT-PCR at all the time points. However, the same for Omicron variant was about 70% lower as compared to the other two lineages at all the time-points (Fig. 3A). Similarly, viral titers in the supernatant increased from 12 h pi to 24 h pi and the levels were maintained at 36 h pi in the case of B.6 lineage virus. However, consistent with the sub-genomic N RNA levels, the viral titers for the Omicron variant was significantly lower compared to both B.6 and the Delta variant at all the time points (Fig. 3B) which is in agreement with recent reports(30, 31). These observations were further corroborated by western blot analysis performed with cell lysates prepared from infected cells collected in lysis buffer at 12 and 24 h pi where the N protein was consistently lower at both 12 and 24 h pi in cells infected with Omicron as compared to B.6 and Delta infection (Fig. 3C). To further verify whether the lower titers observed in the Omicron infection is due to compromised viral entry, we performed viral entry experiments by infecting cells with 5 MOI for one hour and estimated the amount of internalized RNA by qRT-PCR. We found that, contrary to reduced viral titers and viral RNA levels in Omicron infection, the intracellular viral RNA levels were higher as compared to B.6 and the Delta variant suggesting that the reduced viral titers and viral RNA levels is not due to compromised viral entry into these cells (Fig. 3D).

**Figure 3:**
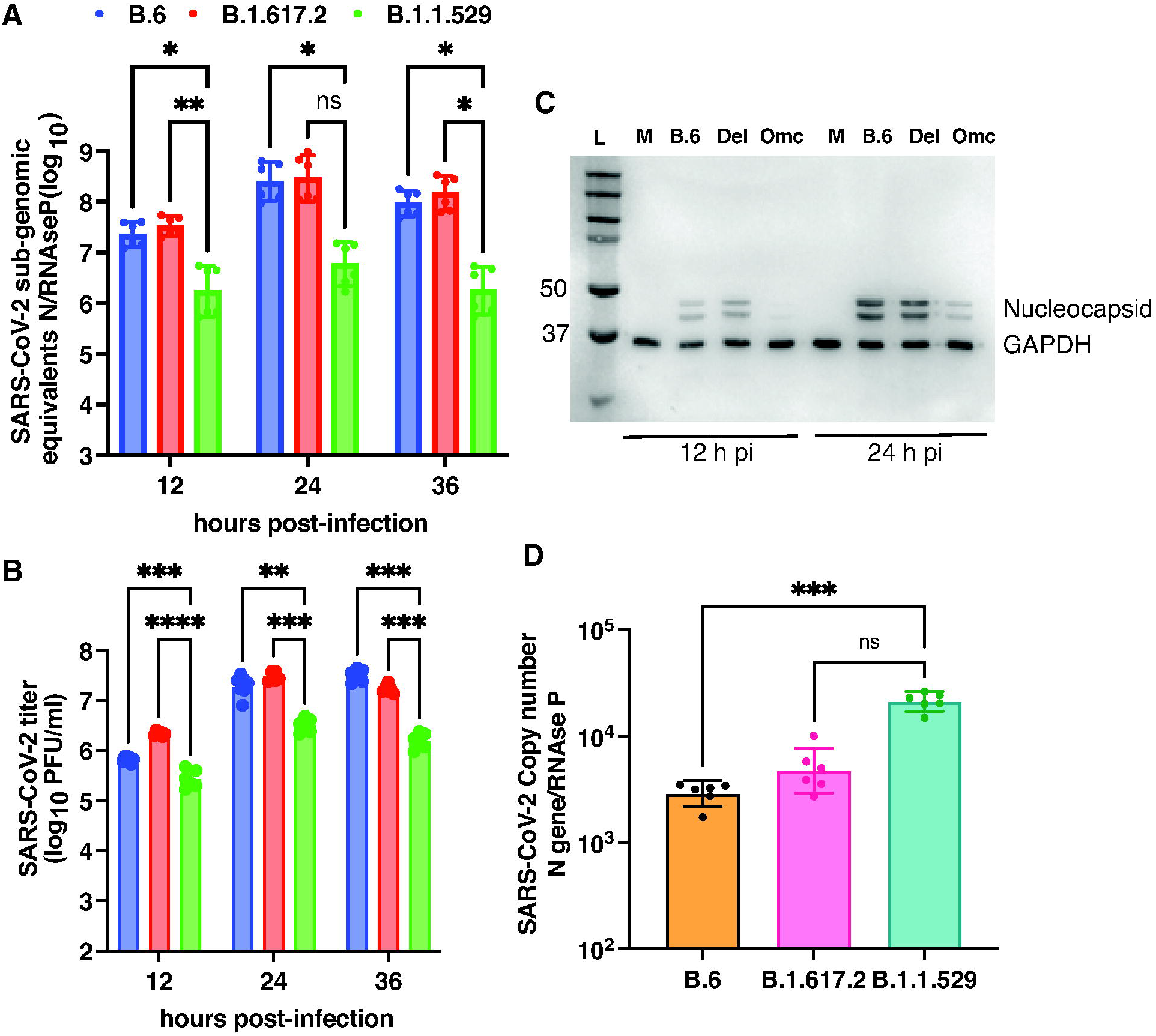
Growth kinetics of B6, B.1.617.2, and B.1.1.529 variants in Calu-3 cells. (A) Calu-3 cells were infected with above variants of SARS-CoV-2 at 0.3 MOI. Cells were collected at indicated time points and total RNA was isolated to estimate *N* gene copy numbers by RT-qPCR. *RNase P* was used as a housekeeping control for normalization. Error bars represent geometric mean with 95% CI. (B) Viral titers were measured in supernatants by plaque assay. Error bars represent geometric mean with 95% CI. Statistical significance was estimated by two-way ANOVA with Dunnett’s multiple comparisons test. (C) Western blot analysis of cell lysates prepared from infected cells at indicated time points to detect the expression of SARS-CoV-2 nucleocapsid. GAPDH was used as a loading control. Numbers on the left indicate the size of the bands in molecular weight marker. (D) Calu-3 cells were infected at 5 MOI and the amount of internalized RNA was estimated by RT-qPCR as described above. Data are from two independent experiments. Error bars represent geometric mean with 95% CI. Statistical significance was estimated by Kruskal-Wallis test with Dunn’s multiple comparison test. ns: non-significant, * P<0.05, ** P<0.01, *** P<0.001, **** P<0.0001.

### Interferon-dependent responses and reduced barrier disruption by Omicron variant

Innate immune responses mediated by interferons act as the first line of defence against viral infections and SARS-CoV-2 was found to be less capable of evading the interferon response compared to SARS-CoV-1(32) (33). Additionally, interferon induction coincided with viral clearance in young adults with mild SARS-CoV-2 infection suggesting a crucial role for antiviral response in resolving infection(34). We tested the induction of interferon-dependent antiviral response genes in cells infected with B.6, Delta and Omicron lineages. Total RNA isolated from Calu-3 cells after indicated time-points post-infection were used to estimate the transcript levels of interferon pathway genes by RT-qPCR (Fig. 4). We found that infection with Omicron variant led to significantly higher induction of interferon-β (Fig. 4A) and the epithelial cell specific interferon-λ1 (Fig. 4B) and the downstream effectors of interferon pathway namely, interferon-stimulated gene 15 (ISG15) (Fig. 4C) and 2’-5’-oligoadenylate synthetase 1 (OAS1) (Fig. 4D) at all the time-points tested suggesting that SARS-CoV-2 ancestral virus (B.6) and both the variants induce innate immune responses and lower levels of titers observed in the Omicron infected cells is due to robust induction of interferon pathway which is in agreement with previous reports(35). To further confirm that the lower levels of replication and infection by Omicron variant translates to compromised ability to disrupt the epithelial barrier functions, we compared the effect of Delta and Omicron variant infection in an air-liquid culture model using Calu-3 cells. Cells were cultured on transwells at the air-liquid interface and infected with respective virus strains at 0.3 MOI. Trans-epithelial electrical resistance was monitored for 36 h (Fig. 5A). We observed a gradual insignificant decline in the TEER values after 12 h pi in cells infected with the Omicron variant and, by 36 h pi, barrier integrity was compromised by 50% relative to mock-infected cells. However, infection with Delta variant led to drastic reduction of >80% in TEER values after 24 h pi suggesting rapid induction of cell death and disruption of epithelial barrier (Fig. 5A). We measured viral titers in the basolateral chamber and, surprisingly, the viral titers were comparable between the Omicron and Delta variant suggesting comparable transcytosis of both these variants but significantly higher cytopathic effect of the Delta variant as compared to the Omicron variant (Fig. 5B). Disruption of cellular junctions was further confirmed by staining the cells with antibodies against SARS-CoV-2 N, occludin and β-catenin. As expected from the TEER values, infection with the Delta variant showed large syncytia formation as visualized by N-positive cells and disruption of and reduction in occludin and β-catenin staining (Fig. 5C). Cells infected with the Omicron variant showed fewer infected cells, mild perturbation in junction proteins (Fig. 5C) further confirming the milder effect of this variant on epithelial barrier functions and pathogenicity as observed in animal studies by other groups(17, 29).

**Figure 4:**
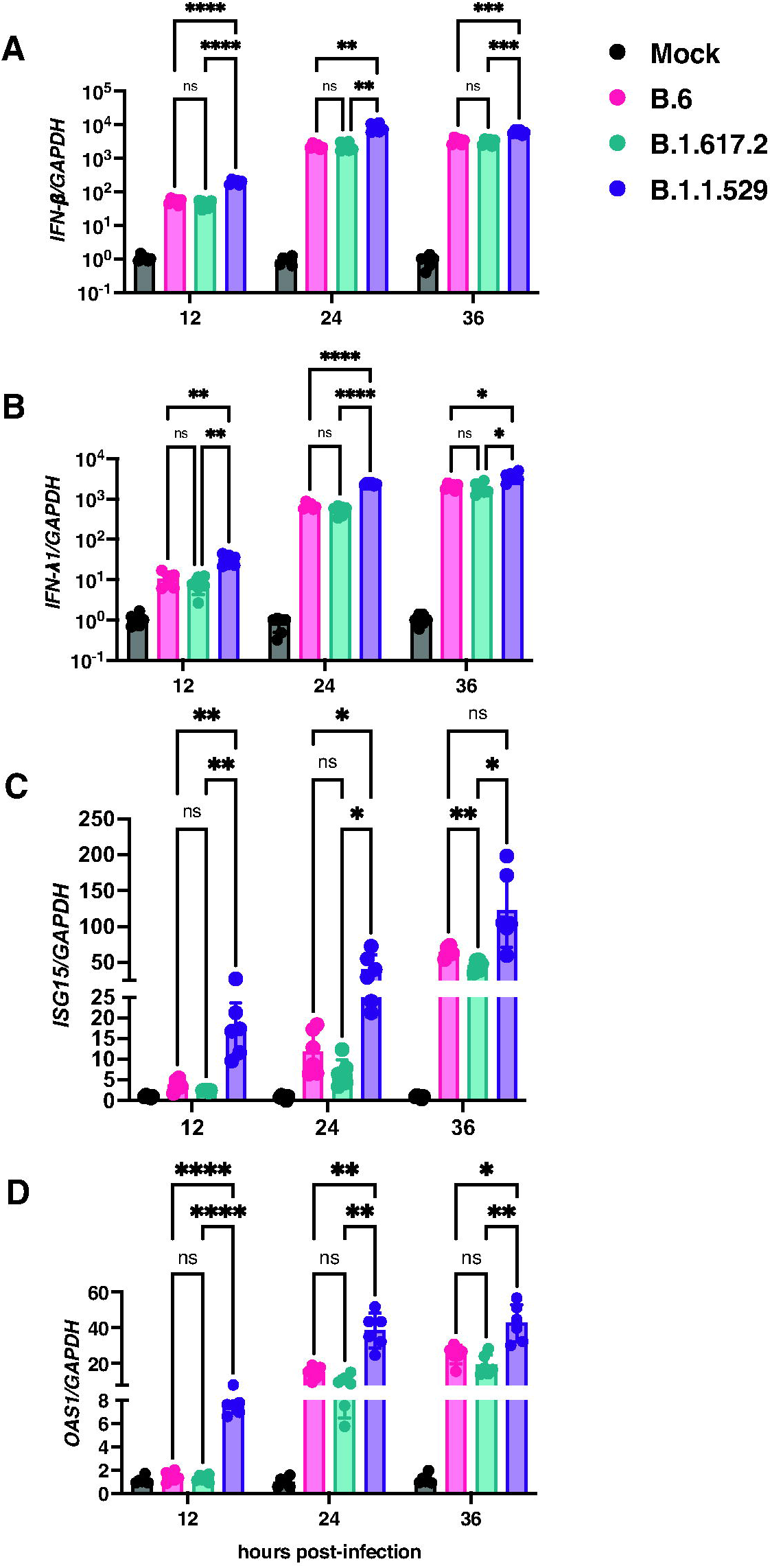
Antiviral response with B.6, B.1.617.2, and B.1.1.529 variants. Calu-3 cells were infected with indicated variants of SARS-CoV-2 at 0.3 MOI. Cells were collected at indicated time-points and total RNA was isolated. RT-qPCR was set up to determine the expression of (A) *IFN-*β. (b) *IFN-*λ*1*, (C) *ISG-15*, and (D) *OAS1. GAPDH* was used as house-keeping control for normalization. Data are from two independent experiments (Mean ± SD). Statistical significance was estimated by two-way ANOVA with Tukey’s multiple comparisons test. ns: non-significant, * P<0.05, ** P<0.01, *** P<0.001, **** P<0.0001.

**Figure 5:**
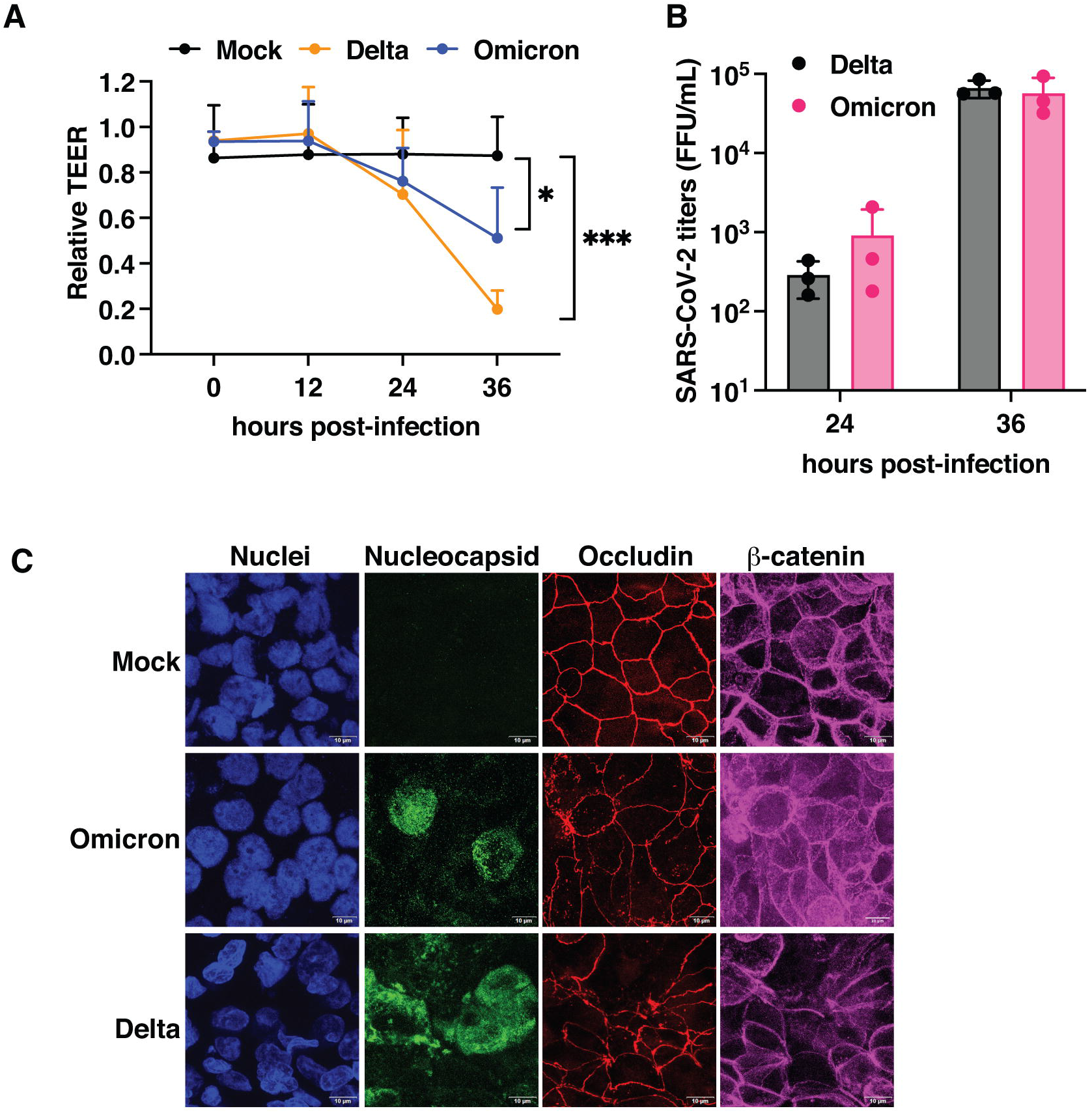
Omicron variant has milder effect on epithelial junctions. Calu-3 cells were grown on transwell inserts under air-liquid interface (ALI) conditions. Cells were infected with Delta and Omicron variants at 0.3 MOI. (A) Graph indicates TEER values relative to mock infection after infection at indicated time points from two independent experiments. (Mean and error with range). Statistical significance was estimated by two-way ANOVA with Tukey’s multiple comparisons test. (B) Viral titers were measured in supernatants by focus-forming units. Error bars represent (Mean ± SD) (C) At 36 h pi, cells were fixed and stained with occludin, β-catenin and SARS-CoV-2 nucleocapsid antibody followed by Alexa Fluor dye-conjugated secondary antibodies for visualization. Nuclei were stained with DAPI. Images were captured at 100X magnification. Images were analyzed using cellSens software and Z-projection images with maximum intensity are shown in the figure. Scale bar is 10 µM. ns: non-significant, **** P<0.0001.

### Comparable replication kinetics and neutralization of BA.1 and BA.2 sub-lineages

Three sub-lineages of Omicron have been identified so far(8, 36) and BA.1 and BA.2 sub-lineages are the circulating virus strains in the third wave in India since November 2021 (Supplementary Fig. S2). We isolated both BA.1 and BA.2 sub-lineage viruses from patient samples and performed growth curve analysis of both these sub-lineages as described earlier in Calu-3 cells. We found no difference in the cellular viral RNA levels and viral titers in the supernatants at the indicated time-points post-infection (Fig. 6A and 6B). To further verify the potential of antibodies from vaccinated individuals to neutralize both BA.1 and BA.2 lineages, we enrolled 15 subjects (Median age 29; 5 Female, 10 Male) who were vaccinated with ChAdOx1 nCoV-19 vaccine in May 2021. We collected samples at three time points: i) first sample at one month after complete vaccination (June 2021), ii) a follow-up sample after 6 months (Dec 2021) and iii) a third sample during the ongoing third wave of SARS-CoV-2 infection (Feb 2022). We performed quantitative nucleocapsid (N) and RBD ELISA(13) to monitor antibody levels in samples from the three bleeds. Six of the 15 samples had detectable levels of N antibodies (cut off: 15 BAU/mL) post-vaccination suggesting that these subjects had hybrid immunity (infection + vaccination) as the ChAdOx1 nCoV-19 vaccine does not generate N antibodies. The geometric mean titer (GMT) of N-ELISA antibodies reduced from 14.0 (95% CI: 8.4, 23.1) in June 2021 to 11.5 (95% CI: 7.9, 16.6) in Dec 2021 with only five positive samples. However, the samples collected during the Omicron surge in Feb 2022 showed a significant increase in N antibodies with a GMT of 53.5 (95% CI: 19.6, 145.8) with ten positive samples (Fig. 6C). Similar trends were observed with RBD antibodies where we observed an approximate 3-fold in the GMT of RBD antibodies from 266.0 (95% CI: 120.2, 368.1) in June 2021 to 76.5 (95% CI: 36.7, 159.5) in Dec 2021. The samples from Omicron wave showed a spike in RBD antibody titers to 935.3 (95% CI: 440.4, 1986) (Fig. 6D).

**Figure 6:**
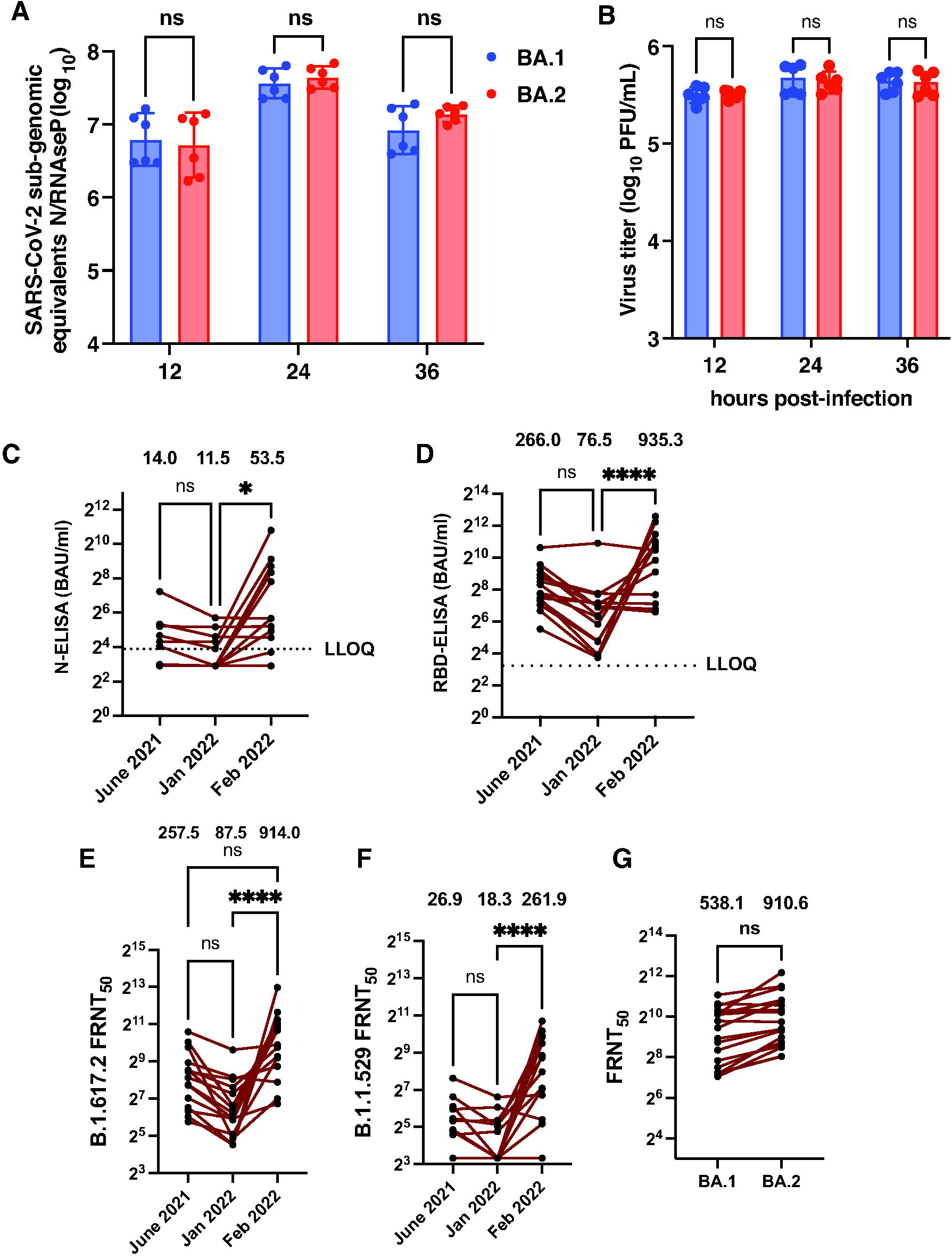
Omicron infection induces cross-protective antibodies: Calu-3 cells were infected with BA.1 and BA.2 sub-lineages of the Omicron variant at 0.3 MOI. Cells were collected at indicated time points and RNA was isolated. (A) SARS-CoV-2 N sub-genomic copy numbers were estimated using RT-qPCR. *RNase P* was used as a housekeeping control. (B) Viral titers were measured in supernatants by plaque assay. Error bars represent geometric mean value with 95% CI (C) Binding antibodies to nucleocapsid (N) and (D) RBD were estimated by quantitative ELISA for the three time-points post-vaccination as indicated (n=15). (E) Neutralizing antibody titers to Delta variant and (F) Omicron variant was determined in the same samples by FRNT assay. (G) Neutralizing antibody titers to the BA.1 and BA.2 sub-lineages were determined in samples from participants that were RT-PCR positive during the ongoing Omicron-induced surge in COVID-19 cases (n=19). Statistical significance was estimated by Kruskal-Wallis test with Dunn’s multiple comparison test. ns: not significant. * P<0.05, **** P<0.0001.

To establish virus neutralization titer assays for the Omicron variant, we first performed conventional plaque assays with the virus stocks using both Calu-3 and Vero E6 cells and found delayed and small plaque formation in Calu-3 cells as compared to the Delta variant (Supplementary Fig. S3A). However, unlike the Delta variant, the Omicron variant did not produce plaques up to 48 h post-infection in Vero E6 cells. As Calu-3 cells are slow-growing and plaque assays are not amenable for high-throughput neutralization assays, we first tested the ability of the Omicron variant to form infectious foci by antibody staining with anti-spike and anti-nucleocapsid antibodies in Vero E6 cells. While the Delta variant formed clear discernable infectious foci with both the antibodies, the same was less distinct with anti-spike antibodies for the Omicron variant although it was countable by the spot reader (Supplementary Fig. S3B). Staining with nucleocapsid antibodies produced better foci in the case of Omicron variant but the quality of foci was not comparable with the Delta variant (Supplementary Fig. S3B). We next tested secondary antibodies conjugated with fluorescent dye instead of horseradish peroxidase enzyme (HRP) and visualized the infectious foci in fluorescence mode. Use of fluorescence-based detection was far superior to HRP-based enzyme-substrate detection for the Omicron variant (Supplementary Fig. S3C). The assay showed consistency in performance as measured by the antibody titers of an in-house pooled convalescent reference serum (Supplementary Fig. S3D). We adopted this method for comparing the neutralizing antibody titers against the Delta and Omicron variants. The neutralizing antibody titers for Delta variant as measured by FRNT assay also showed about three-fold reduction from a GMT of 257.5 (95% CI: 145.8, 454.7) in June 2021 to 87.5 (49.7, 154.1) in Dec 2021 (Fig.6E). However, as expected from the ELISA titers, the GMT of neutralizing antibodies increased significantly to 914.0 (95% CI: 468.7, 1782) in samples from the third wave in Feb 2022 indicating re-infection in most of these individuals. As shown by us and others(10, 37), the neutralizing antibody titers for Omicron variant was drastically reduced at all the time points relative to that of Delta variant (Fig. 6F). Nine out of 15 samples had FRNT_50_ value above the level of detection after vaccination in June 2021. The GMT of neutralizing antibodies for the Omicron variant was 26.9 (95% CI: 15.6, 46.4) in these samples. The number of samples positive for Omicron antibodies reduced to six out of 15 bringing the down the GMT of neutralizing antibodies to 18.3 (95% CI: 11.6, 28.7) by Dec 2021. As expected, the surge in Omicron infections in the study area was reflected in the GMT of neutralizing antibodies in samples from Feb 2022 (Fig. 6F). Surprisingly, the samples from Omicron surge showed relatively lower titer, a GMT of 261.9 (95% CI: 113.0, 607.5) as compared to a GMT of 914.0 (95% CI: 468.7, 1782) for the Delta variant suggesting that natural infection with Omicron variant generates significantly higher levels of antibodies to the Delta variant.

SARS-CoV-2 sequencing data from the National Capital Region (NCR), our study site, showed co-occurrence of B.1.617.2 (Delta) and various AY.* (Delta plus) lineages during the months of September 2021 to December 2021, wherein all AY lineages were more in circulation than the parent B.1.617.2 lineage (Supplementary Figure S2). It is important to note that a rise in the cases of Omicron lineages (B.1.529, BA.1 and BA.2) is observed from December 2021 onwards. This is concomitant with the arrival of Omicron variant in India and a gradual decrease in circulation of Delta lineage. Delta variant was superseded by Omicron by January 2022. Within Omicron lineages, cases of BA.2 sub-lineage increased/spread at a faster rate compared to other Omicron lineages (BA.1 and B.1.529) (Supplementary Figure S2) which is similar to reports from other countries. This is consistent with the observations that BA.2 lineage has increased rates of transmission (38, 39). We enrolled participants (n=19) who were either COVID-19 RT-PCR positive in the ongoing Omicron surge or had symptomatic respiratory infection but got no COVID-19 testing done. The GMT of neutralizing antibodies for BA.1 and BA.2 sub-lineages in these samples were 538.1 (95% CI: 338.5, 855.3) and 910.6 (95% CI: 608.2, 1363) and this difference was not statistically significant as reported by others(40). As we recently reported minimal to no neutralizing antibodies against the Omicron variant in subjects with vaccine-induced or hybrid immunity at the beginning of the surge in COVID-19 cases in December 2021(12), our data suggests that enhanced neutralizing antibody titers observed in this study cohort is due to natural exposure to SARS-CoV-2 Omicron variant.

### Copper and Iron (II) salts inhibit both Delta and Omicron variant infection

Divalent cations play an important role in replication of RNA viruses(41) and previous reports have suggested inhibition of coronavirus replication by Zn salts(42). We established the fluorescence-based SARS-CoV-2 RNA-dependent RNA polymerase (RdRp) assay using purified nsp7, nsp8 and nsp12 proteins with minor modifications based on previous reports(43-45). RdRp assays were performed in the presence of 10 μM, 100 μM or 1000 μM of CaCl_2_/CuCl_2_/FeSO_4_/ZnSO_4_. CaCl_2_ did not affect RdRp activity under these conditions. At 1000 μM, CuCl_2_ showed about 35% reduction in RdRp activity while FeSO_4_ and ZnSO_4_ showed 85% and 60% inhibition respectively at the same concentration (Fig. 7A). At 100 μM, FeSO_4_ showed 25% reduction in activity which was significant compared to untreated while ZnSO_4_ showed 52% inhibition at this concentration (Fig. 7A). Only ZnSO_4_ was capable of inhibiting SARS-CoV-2 RdRp activity albeit by only 22% at 10 μM (Fig. 7A). The 50% inhibitory concentration (IC_50_) of ZnSO_4_ and FeSO_4_ in RdRp assays was 313 μM and 347 μM respectively (Fig. 7B and 7C). To further confirm the validity of these *in vitro* inhibition in cell culture, we infected Calu-3 cells with either the Delta variant or the Omicron variant and medium supplemented with 50 μM each of CaCl_2_/CuCl_2_/FeSO_4_/MnCl_2_/ZnSO_4_ was added after virus adsorption and cells were cultured for 24 h. This concentration of salts was shown to not affect viability of the cells (Supplementary Figure S4). Virus titers in the infected culture supernatants were estimated by plaque assays. Contrary to RdRp assay results, ZnSO_4_ at 50 μM concentrations failed to show any inhibition of either the Delta or the Omicron variants in Calu-3 cells (Fig. 7D and 7E). Similarly, neither CaCl_2_ nor MnCl_2_ had any impact on virus titers. However, both CuCl_2_ and FeSO_4_ showed significant inhibition of virus titers. Both these salts inhibited the Delta variant infection by 70% (Fig. 7D) and the effect was even more drastic (>90% inhibition) in the case of Omicron variant (Fig. 7E). Therefore, our results suggest that copper and iron salts may have both direct and indirect antiviral activity against SARS-CoV-2 when added post-infection.

**Figure 7:**
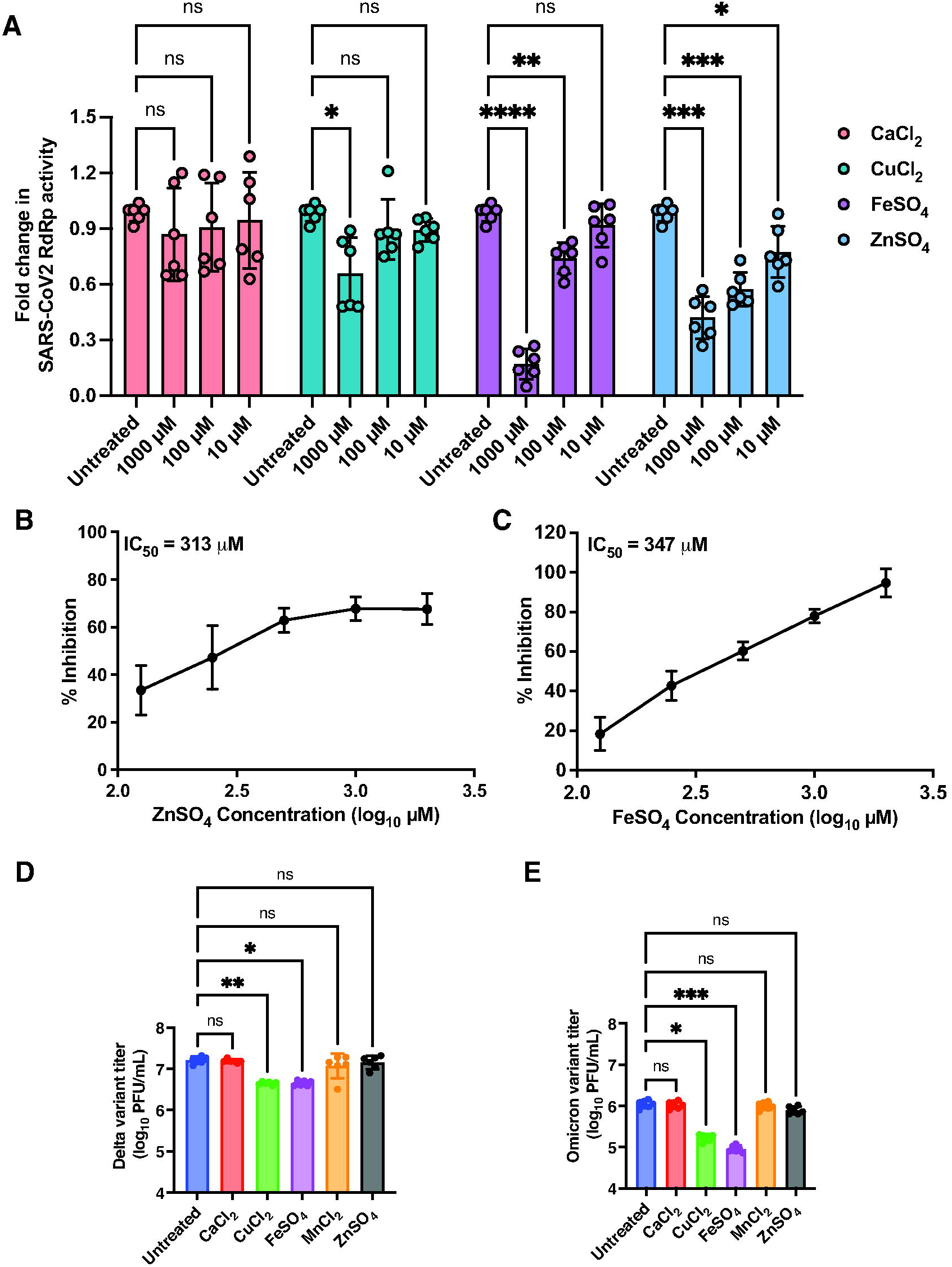
Iron and copper salts significantly inhibits SARS-CoV-2 infection. (A) SARS-CoV-2 RdRp activity assay was performed in the presence of indicated concentrations of CaCl_2_/CuCl_2_/FeSO_4_/ZnSO_4_ in the standard reaction mix containing MgCl_2_ and MnCl_2_ and the RdRp complex (nsp7-nsp8-nsp12) of SARS-CoV-2. (B) RdRp assays were performed in the presence of varying of concentrations of ZnSO_4_ or (C) FeSO_4_ to determine the IC_50_ values. To determine the effect of metal ions in infection, Calu-3 cells were infected with indicated variants at 0.3 MOI and media containing 50 µM each of the indicated salts were added after virus adsorption. At 24 h pi, supernatants were collected and viral titers were measured by plaque assay. The graph indicates the virus titers of (D) Delta and (E) Omicron variants. Data are from two or three independent experiments. Error bars represent geometric mean with 95% CI. Statistical significance was estimated by Kruskal-Wallis test with Dunn’s multiple comparison test. ns: non-significant, * P<0.05, ** P<0.01, *** P<0.001, **** P<0.0001.

## DISCUSSION

SARS-CoV-2 Delta variant was shown to be more infectious and duration from PCR positivity or symptoms with this variant was shorter as compared to other SARS-CoV-2 virus strains leading to faster transmission and severe disease. In our study, the Delta variant showed enhanced disruption of epithelial barriers and junction proteins suggesting that the enhanced cytopathic effect and syncytia formation combined with the ability to escape from neutralizing antibodies has provided a growth advantage for Delta variant(1, 2, 28). Contrary to the enhanced pathogenicity of the Delta variant, the Omicron variant showed lower replication and had milder effect on epithelial barrier functions which correlates with less number of severe cases, hospitalization and lower number of deaths due to COVID-19 in the ongoing third wave of SARS-CoV-2 infections in India which is similar to what has been reported from other countries(46). The clinical samples for this study was collected during the second and third wave of COVID-19 in India during which the Delta and Omicron variants were the predominant circulating virus strains. Cohort data of patients with mild COVID-19 infection has shown that RBD antibodies decay with a half-life of 69 days(47). We observed a three-fold reduction in RBD antibody and neutralizing antibody titers after six months. However, most of the samples had no neutralizing antibodies against the Omicron variant after six months of vaccination suggesting that waning neutralizing titers are an important determinant of reinfections and may also contribute to increased susceptibility to new variants of concern that escape pre-existing humoral immunity. The high-throughput neutralizing antibody assay we established in this study enabled us to circumvent the challenges of estimating neutralizing antibody titers for the Omicron variant as it grew poorly and did not form distinct plaques or infectious foci like the Delta variant. Currently, BA.1 and BA.2 sub-lineages of the Omicron variant are circulating in India and reports have raised concern about the enhanced pathogenicity or transmissibility of BA.2 sub-lineage due to unique mutations in the spike region(39, 48). We show that both BA.1 and BA.2 sub-lineages of Omicron variant have comparable replication kinetics in Calu-3 cells. Our sequencing data suggests that the BA.2 sub-lineage of the Omicron variant was the predominant circulating virus at the time of the study and relatively higher levels of neutralizing antibodies to BA.2 was observed in study participants as compared to BA.1 further confirming that BA.2 may have been the infecting virus. We show that participants who were naturally infected with the Omicron variant generated neutralizing antibodies against both BA.1 and BA.2 sub-lineages indicating that exposure to any of the sub-lineages of Omicron would confer cross-protection against other members of the sub-lineage in agreement with other reports(14, 40). Interestingly, higher titers of neutralizing antibodies were observed against the Delta variant which was no longer in circulation indicating a broader cross-protective immune response most likely due to conserved B-cell and T-cell epitopes in the Omicron variant that could have triggered the memory responses in people with hybrid immunity(49, 50). Thus, natural infection with Omicron variant may have acted as a booster eliciting cross-reacting antibodies across Omicron sub-lineages and other VoCs in young adults.

Several studies have reported that interferons expressed in the lower respiratory tract contribute to tissue damage, impair regeneration of lung epithelium and contribute to morbidity observed in COVID-19 patients(51, 52). We observed similar levels of induction of *IFN-β* and *IFN-λ1* and the downstream effectors namely *ISG15* and *OAS1* in cells infected with the Delta variant and with the ancestral B.6 lineage virus suggesting that suppression of innate immune responses is not a contributory factor in increased pathogenesis observed with the Delta variant. Despite lower levels of infection, the Omicron variant induced higher levels of interferon pathway genes. Since the Omicron variant was shown to mostly replicate in the upper respiratory tract(53, 54), this suggests that interferon-dependent antiviral responses may limit viral replication and further tissue spread thus resulting in milder symptoms and resolution of infection. Therefore, interferon therapy targeted towards the upper respiratory tract may be more effective in the case of infection with the Omicron variant. In addition to interferons, our results suggest that supplementation with copper and iron salts may also augment the antiviral strategy for SARS-CoV-2 as both CuCl_2_ and FeSO_4_ inhibited both the Delta and Omicron variants in Calu-3 cells.

Vaccines have played a major role in bringing the COVID-19 pandemic under control. However frequent emergence of SARS-CoV-2 variants requires adjunct approaches to deal with infections. Micronutrient supplementation could be an effective and safe strategy. Ferric ammonium citrate (FAC) was shown to inhibit a number of RNA viruses including Influenza A virus, Zika virus and Hepatitis C virus in both cell culture and animal models and it was shown that treatment with FAC perturbed endosomal release of the internalized virus in the case of Influenza A virus(55). SARS-CoV-2 nsp12 has been shown to bind Fe and Iron-Sulfur cluster binding was required for optimal activity of the RdRp(56). However, there have been no direct demonstration of the effect of Fe salts on either the RdRp activity in vitro on in lung epithelial cells. We show here that FeSO_4_ inhibited both RdRp activity and infection in Calu-3 cells. The inhibitory concentration in cell culture was much below the *in vitro* IC_50_ concentrations. Contrary to our findings, iron overload is considered as a risk factor for severe COVID-19 disease and iron chelation has been proposed as a treatment option(57) (58). Therefore, the exact mechanism of action behind inhibition of SARS-CoV-2 infection by FeSO_4_ warrants further investigation. Interestingly, CuCl_2_ which inhibited RdRp activity at 1 mM also inhibited virus infection in Calu-3 cells at 20 times below this concentration indicating that both Fe an Cu ions may act on SARS-CoV-2 both directly and via host pathways. Previous reports have shown that copper-coated surfaces can inactivate SARS-CoV-2(59) and other enveloped and non-enveloped viruses are susceptible to copper ions either due to direct effect of copper ions on viral particles or viral genome or due to induction of reactive oxygen species. Therefore, copper supplementation during viremic phase may further augment antiviral strategies for COVID-19(60-62). Zinc acetate, in combination with zinc ionophore pyrithione, was shown to inhibit SARS-CoV-1 infection in Vero E6 cells and zinc ions was shown to directly inhibit RdRp activity of SARS-CoV-1(42). SARS-CoV-2 proteome encodes for a number of metalloproteins including zinc-binding proteins such as nsp2, nsp9, nsp10/16 complex, nsp12 and proofreading exoribonuclease nsp14 (Please see Uniprot ID: P0DTD1 · R1AB_SARS2). Some of the critical aspects of viral replication such as RdRp activity, 2’-*O* methylation, capping depends on intracellular zinc(63, 64). Zinc ejection from some of these enzymes has been proposed as an attractive antiviral target(65). However, despite ZnSO_4_ inhibiting the activity of SARS-CoV-2 RdRp *in vitro*, we saw no effect on SARS-CoV-2 Delta and Omicron variants at 50 μM concentration. Whether higher concentrations of zinc salts are required to achieve significant inhibition of these variants in cell culture needs further investigation.

## ONLINE METHODS

### Human Ethics

The study was approved by the Institutional Ethics Committee for human research at ESIC Hospital and Medical College (No.134/R/10/IEC/22/2021/02) and THSTI (THS 1.8.1/ (93)). Informed consent was obtained from all the participants.

### Human Samples

ChAdOx1 nCoV-19 cohort: Subjects (age 25-46) visiting ESIC Medical College & Hospital, Faridabad for vaccination were enrolled in the study after obtaining written informed consent. About 4 ml of whole blood was collected for serum preparation four weeks after the first and second dose of vaccination and a follow-up sample was collected after six months. Nasopharyngeal/Oropharyngeal (NP/OP) swabs were collected from patients with symptoms of COVID-19 infection. Total RNA was isolated to detect SARS-CoV-2 using COVIDsure multiplex real-time RT-PCR kit (Trivitron Healthcare) either at Employees State Insurance Corporation (ESIC) Medical College & Hospital or at the bioassay laboratory Translational Health Science and Technology Institute, Faridabad. Clinical presentations were mild to moderate fever, dry cough, and loss of sense of smell and taste. All COVID-19 positive patients were self-isolated and recovered without any need for clinical intervention or hospitalization. A follow-up blood sample was collected after 3-4 weeks post-recovery in both second (May 2021) and third wave (Feb 2022).

### Cells and Viruses

Calu-3 cells and Vero E6 cells were procured from American Type Culture Collection (ATCC) and European Collection of Authenticated Cell Cultures (ECACC) respectively. Both cell lines were maintained in Dulbecco’s Modified Eagle Medium (DMEM) (HiMedia) supplemented with 10% fetal bovine serum (FBS), 100 units/ml of Penicillin-Streptomycin-Glutamine (PSG), and 1X non-essential amino acid (NEAA). In addition to this, 25 mM HEPES was added to the Vero E6 culture medium. Different variants of SARS-CoV-2 i.e., ancestral (B.6, Genbank: MW422884.1), kappa (B.1.617.1, Genbank: MZ356902.1), delta (B.1.617.2, Genbank: MZ356566.1), and omicron (B.1.529; BA.1 GISAID: EPI_ISL_6716902 and BA.2 GISAID ID: EPI_ISL_87638432 sub-lineages) were propagated in Calu-3 cells and passaging was limited to five passages.

### Growth curve experiment

All experiments with infectious SARS-CoV-2 virus were performed at the infectious disease research facility, which is a biosafety level-3 laboratory. Calu-3 cells were seeded in 48-well or 24-well plates and infected with indicated SARS-CoV-2 virus isolates two days post-seeding. Infected culture supernatants were collected at indicated time points to estimate viral titers by plaque assay on Vero E6 cells. Total RNA was extracted from cells collected at respective time points and viral copy numbers (N gene) were estimated by quantitative RT-PCR using RNAse P gene for normalization as described previously(66).

### Plaque assay Method

Vero E6 cells were used for virus titration by plaque assay method. The supernatants were collected from infected cells at indicated time points. The supernatants were ten-fold diluted using growth medium with 2% FBS. After 1h of viral adsorption, virus inoculum was removed. Overlay medium with 0.5% carboxymethylcellulose (CMC) was added to the cells. The plates were incubated at 37° C for 48 h for all SARS-CoV-2 variants except omicron where the incubation period was 72 h. After the incubation period, cells were fixed with 3.7% formaldehyde solution followed by incubation for 10 min at room temperature (RT). Cells were stained with crystal violet solution and plaques were observed.

### Trans-epithelial electrical resistance (TEER) and confocal microscopy

Calu-3 cells were seeded at 30,000 on 3 µm pore size transwell inserts (Corning - 3415) and grown for 21 days. TEER was monitored on alternate days using a chopstick electrode (Millipore) and the medium was changed on the same day. On day 22 post-seeding, cells were infected with indicated variants of SARS-CoV-2 at 0.3 MOI. After 1 h of viral adsorption, virus inoculum was removed and cells were washed twice with 1X PBS. Growth medium with 10% FBS was added to the cells. TEER was monitored at indicated time points. At 48 h pi, culture inserts were washed with cold 1X PBS and incubated with cold methanol at −20°C for 20 minutes. Cells were further incubated with IMF buffer (20 mM HEPES, pH 7.5, 0.1% Triton X-100, 150 mM sodium chloride, 5 mM EDTA and 0.02% sodium azide as a preservative) for 5 min at room temperature (RT) and all further washes were performed with IMF buffer. Non-specific antibody binding sites were blocked by incubating cells with IMF buffer containing 2% normal goat serum for 10 min at RT. Membranes were cut out using a scalpel blade and transferred to a 48-well plate. Cells were washed three times followed by incubation with antibodies against SARS-CoV-2 nucleocapsid, occludin, and β-catenin in IMF buffer for 1 h at RT. Cells were washed followed by incubation with secondary antibodies tagged with Alexa Fluor 488/568/633 (Molecular probes) for 30 min at RT by avoiding exposure to light. Cells were washed with IMF buffer three times and stained with DAPI at 1:10,000 dilution for 10 min. Cells were washed with PBS, mounted on glass slide using antifade solution (Molecular probes), and images were captured at 100X magnification using FV3000 confocal microscope (Olympus). Acquired images were processed using CellSens software (Olympus) and projections of Z-stack (maximum intensity) are shown in figures.

### Focus-forming units (FFU) assay

The supernatants were collected from infected cells at indicated time points. The supernatants were ten-fold diluted using growth medium with 2% FBS. After 1h of viral adsorption, virus inoculum was removed. Overlay medium with 1.5% carboxymethylcellulose (CMC) for all SARS-CoV-2 variants except for Omicron where 1% CMC was used. The plates were incubated at 37° C for 28 h for omicron and 24 h for all other SARS-CoV-2 variants. After incubation time, cells were fixed with formaldehyde solution followed by permeabilization with IMF buffer for 20 min incubation. Further, cells were stained with anti-spike RBD rabbit polyclonal antibody dilution at a dilution of 1:2000 for 1 h, followed by incubation with secondary antibody i.e., Alexa flour 488-conjugated anti-rabbit antibody at 1:500 dilution for 1 h. For omicron isolate, anti-nucleocapsid primary antibody was used at a dilution of 1:2000. This was followed by incubation with secondary antibody i.e., Alexa flour 488-conjugated goat anti-mouse IgG secondary antibody at 1:500 dilution. Fluorescent foci indicating infected cells were observed and counted using AIDiSpot reader using FITC channel.

### Western blot

At indicated time points, cells were washed twice with cold PBS on ice and were collected in 200 µl of 1X laemmli buffer with protease inhibitor mix and 1 mM PMSF. Lysates were incubated on ice for 10 min and sonicated (amplitude: 30%, time: 5 sec ON/10 sec OFF, total cycles: 10) on ice. Cell lysates were centrifuged at 13,000 x *g* for 15 min at 4 °C. 5 µl of each sample was resolved on 12% SDS-PAGE. Gels were transferred onto PVDF membrane for 2 h and the SARS-CoV-2 infection was analyzed by probing the blot with the SARS-CoV-2 nucleocapsid antibody. GAPDH antibody was used as a loading control. Primary antibody incubation was followed by HRP-conjugated secondary antibodies. Luminol-based chemiluminescent substrate was added to the blots and signals were detected using a gel documentation system (Azure biosystems C400).

### Virus entry experiment

Calu-3 cells were infected with the above-indicated variants of SARS-CoV-2 at 5 MOI. After 1 h of viral adsorption, the virus inoculum was removed and cells were washed with 1X PBS. Cells were trypsinized and collected in complete growth medium and centrifuged at 200 x *g* for 5 min. The supernatants were discarded and the cell pellets were washed twice with 1X PBS followed by centrifugation. In the final step, the cell pellets were collected in 350 µl lysis buffer. Viral copy numbers were estimated as described in the above section.

### Quantitative RT-PCR Assay

Calu-3 cells were infected with ancestral, delta, and omicron variants of SARS-CoV-2 at a MOI of 0.3. At indicated time points, the supernatant was collected for estimating viral titres by plaque assay, and cells were collected in lysis buffer provided in the RNA isolation kit as described in the above section. For interferon pathway, 500 ng RNA was used to erase genomic DNA and reverse transcribed using PrimeScript RT reagent kit with gDNA eraser kit. The indicated genes were quantitated by SYBR green chemistry and GAPDH was used as housekeeping control gene for normalization. The list of primers used is provided in the key resources table. Data were analyzed using the *ΔΔC*_*T*_ method, where *C*_*T*_ is cycle threshold.

### TEER in air-liquid interface condition and confocal microscopy

Calu-3 cells were seeded at a density of 30,000 cells in 3 μm pore size polycarbonate membranes in 24 well-plate with corresponding 200 µl culture medium on the apical side and 800 µl culture medium was added into the basolateral chamber. Cells were left to grow submerged for 8 days. The culture medium was changed on alternate days and TEER values were monitored. On day 9, the medium was removed from the apical chamber to obtain air-liquid interface (ALI) conditions. Cells were cultured in ALI conditions for 20 days. After obtaining stable TEER values, on day 21, cells were washed with 1X Hanks’ balanced salt solution (HBSS). Cells were infected with either the Delta or the Omicron variants of SARS-CoV2 at 0.3 MOI. After 1 h of viral adsorption, cells were washed twice with HBSS, and culture medium was added to the basolateral side. To assess the effect of SARS-CoV-2 on epithelial cells barrier, TEER was monitored at indicated time points. Supernatants were collected from basolateral surface for measuring viral titres. At 36 h pi, cells were washed with cold PBS and fixed in ice-cold methanol for 20 min at −20°C. The cells were stained against junction proteins along with nucleocapsid antibodies as described above in the confocal microscopy section.

### Quantitative Nucleoprotein (N) and RBD ELISA

ELISA for nucleocapsid (N) and recombinant spike protein receptor-binding domain (RBD) was performed as described earlier. Detailed method description is provided in previous reports(13, 67).

### Virus microneutralization assay

Virus microneutralization assay by focus reduction neutralization titer (FRNT) assay using indicating virus isolates was performed as described earlier(13, 67).

### RNA-dependent RNA polymerase (RdRp) activity assay

The Quant-iT™ PicoGreen^®^ dsDNA reagent was used to establish a fluorophore-based RdRp activity system. Adenosine 5′-triphosphate (ATP) was purchased from Sigma–Aldrich. RdRp RNA template with self-hairpin primer used for RdRp assays in this study as following 5′-UUUUUUUUUUUUUUUUUUUUUUUUUUUUUUAACAGGUUCUAGAACCUGUU-3′ (Eurofins). Each reaction (final volume 30 µl) was carried out in 96 well black flat-bottom plates in triplicates. 150 nM of nsp12 and 1.5 µM of nsp7 & 8 was mixed in a reaction buffer (50 mM HEPES pH 8, 5 mM DTT, 10 mM KCl, 2 mM MgCl_2_, 2 mM MnCl_2_) to make up to 15 µl. Indicated concentrations of ZnSO_4_, CaCl_2_, CuCl_2,_ and 5 μl of 200 nM poly-U were added to the reaction buffer. The mixture was incubated at 30 □ C for 5 min. The reactions were initiated by adding 5 μl of 500 µM ATP to the reaction mixture. The plate was incubated at 30 □C for 30 min. The reaction was stopped by adding 20 µl EDTA (100 mM). The Quant-iT™ PicoGreen^®^ dsDNA reagent was diluted to 1/800 in 1X TE buffer (10 mM Tris-HCl, 1 mM EDTA, pH 7.5) and 50 µl of this reagent was added to each well of the plate. The plate was incubated in the dark for 30 min at RT. The fluorescence signal was assessed using microplate reader at excitation of 485 nm and emission of 582 nm.

### Inhibition experiments

Calu-3 cells were seeded at a density of 90,000 per well in 48 well-plate. Cells were infected with indicated SARS-CoV2 variants at 0.3 MOI. After viral adsorption, cells were washed twice with 1X PBS. CaCl_2_, CuCl_2_, FeSO_4_, MnCl_2,_ and ZnSO_4_ were added each in growth medium in serum-free condition to a final concentration of 50 µM. After 24 h pi, supernatants were collected and viral titers were measured by plaque assay.

### Statistical tests

Data were analysed and charts were prepared using GraphPad Prism software. All experiments were performed with two or more replicates and graphs have been prepared representing data from at least two independent experiments. Figure legends indicate error bars and statistical tests conducted for estimating P values.

### Data availability

All the data are presented in this manuscript. Source data will be made available upon request.

## Supporting information

Combined Supplementary data

## Data Availability

All data produced in the present work are contained in the manuscript

## ACKNOWLEDGEMENTS

We thank all the members of bioassay lab, COVID-19 testing team, Amresh Kumar Singh Maniram and other members of the Clinical and Cellular Virology lab for technical support. We thank Neha Garg and Shamsher Singh for data management. We thank all the patients who consented to participate in the study.

## AUTHOR CONTRIBUTIONS

JS, AP, AA, MB, PK and KP performed all cell culture and infection experiments. CR purified recombinant proteins and performed the *in vitro* RdRp assays. PC, PD, RM, PM performed SARS-CoV-2 genome sequencing and analysis. AKP supervised and coordinated the study at the clinical site and contributed reagents. RP designed the sequencing experiments, supervised the team and analyzed the data. GRM conceived the study, designed the experiments, and analyzed the data. JS, AP, AA and GRM wrote the manuscript. All authors have reviewed and approved the final version of the manuscript.

## FUNDING INFORMATION

This work was supported by the Department of Biotechnology (DBT) grants through IndCEPI Mission (BT/MB/CEPI/2016), Translational Research Program (BT/PR30159/MED/15/188/2018) and BT/PR40384/COT/142/5/2020. Funding support was also obtained from Global Immunology and Immune Sequencing for Epidemic Response (INV-030592). RP acknowledges funding support from IUSSTF (CLP-0033) and BMGF (CLP-0036). The funders had no role in study design, data collection and interpretation or the decision to submit the work for publication.

## CONFLICT OF INTEREST STATEMENT

The authors have declared that no conflict of interest exists.

## SUPPLEMENTARY INFORMATION

Supplementary methods and Supplementary figure legends have been provided as a separate file.

## Notes

### Competing Interest Statement

The authors have declared no competing interest.

### Funding Statement

This work was supported by the Department of Biotechnology (DBT) grants through IndCEPI Mission (BT/MB/CEPI/2016) and Translational Research Program (BT/PR30159/MED/15/188/2018). Funding support was also obtained from Global Immunology and Immune Sequencing for Epidemic Response (INV-030592). RP acknowledges funding support from IUSSTF (CLP-0033) and BMGF (CLP-0036). The funders had no role in study design, data collection and interpretation or the decision to submit the work for publication.

### Author Declarations

The study was approved by the Institutional ethics committees for human research at ESIC Hospital and Medical College (No.134/R/10/IEC/22/2021/02) and THSTI (THS 1.8.1/ (93)). Informed consent was obtained from all the participants.

### Summary of Updates

The revised manuscript has all new figures with new data. Figure 3 from previous version is now supplementary figure S3. Previous figure 4 is now part of new figure 6.

## REFERENCES

1. Campbell F, Archer B, Laurenson-Schafer H, Jinnai Y, Konings F, Batra N, et al. Increased transmissibility and global spread of SARS-CoV-2 variants of concern as at June 2021. Euro Surveill. 2021 Jun;26(24). PubMed PMID: 34142653. Pubmed Central PMCID: PMC8212592. Epub 2021/06/19.

2. Fisman DN, Tuite AR. Progressive Increase in Virulence of Novel SARS-CoV-2 Variants in Ontario, Canada. medRxiv. 2021:2021.07.05.21260050.

3. Li B, Deng A, Li K, Hu Y, Li Z, Xiong Q, et al. Viral infection and transmission in a large well-traced outbreak caused by the Delta SARS-CoV-2 variant. medRxiv. 2021:2021.07.07.21260122.

4. Mlcochova P, Kemp S, Dhar MS, Papa G, Meng B, Mishra S, et al. SARS-CoV-2 Delta variant emergence, replication and sensitivity to neutralising antibodies. bioRxiv. 2021:2021.05.08.443253.

5. Bernal JL, Andrews N, Gower C, Gallagher E, Simmons R, Thelwall S, et al. Effectiveness of COVID-19 vaccines against the B.1.617.2 variant. medRxiv. 2021:2021.05.22.21257658.

6. Planas D, Veyer D, Baidaliuk A, Staropoli I, Guivel-Benhassine F, Rajah MM, et al. Reduced sensitivity of infectious SARS-CoV-2 variant B.1.617.2 to monoclonal antibodies and sera from convalescent and vaccinated individuals. bioRxiv. 2021:2021.05.26.445838.

7. Supasa P, Zhou D, Dejnirattisai W, Liu C, Mentzer AJ, Ginn HM, et al. Reduced neutralization of SARS-CoV-2 B.1.1.7 variant by convalescent and vaccine sera. Cell. 2021 Apr 15;184(8):2201–11 e7. PubMed PMID: 33743891. Pubmed Central PMCID: PMC7891044. Epub 2021/03/22.

8. Mahase E. Covid-19: What do we know about omicron sublineages? BMJ. 2022 Feb 11;376:o358. PubMed PMID: 35149516. Epub 20220211.

9. Cele S, Jackson L, Khan K, Khoury DS, Moyo-Gwete T, Tegally H, et al. SARS-CoV-2 Omicron has extensive but incomplete escape of Pfizer BNT162b2 elicited neutralization and requires ACE2 for infection. medRxiv. 2021 Dec 11. PubMed PMID: 34909788. Pubmed Central PMCID: PMC8669855. Epub 20211211.

10. Dejnirattisai W, Shaw RH, Supasa P, Liu C, Stuart AS, Pollard AJ, et al. Reduced neutralisation of SARS-CoV-2 omicron B.1.1.529 variant by post-immunisation serum. Lancet. 2021 Dec 20. PubMed PMID: 34942101. Pubmed Central PMCID: PMC8687667. Epub 20211220.

11. Iketani S, Liu L, Guo Y, Liu L, Huang Y, Wang M, et al. Antibody Evasion Properties of SARS-CoV-2 Omicron Sublineages. bioRxiv. 2022:2022.02.07.479306.

12. Medigeshi GR, Batra G, Murugesan DR, Thiruvengadam R, Chattopadhyay S, Das B, et al. Sub-optimal neutralisation of omicron (B.1.1.529) variant by antibodies induced by vaccine alone or SARS-CoV-2 Infection plus vaccine (hybrid immunity) post 6-months. eBioMedicine. 2022;78.

13. Singh J, Shaman H, Anantharaj A, Singh B, Pargai K, Kumar P, et al. Infection with Omicron variant generates neutralizing antibodies to BA.1 and BA.2 sub-lineages and induces higher levels of cross-neutralizing antibodies to Delta variant. medRxiv. 2022:2022.01.28.22269990.

14. Yu J, Collier AY, Rowe M, Mardas F, Ventura JD, Wan H, et al. Neutralization of the SARS-CoV-2 Omicron BA.1 and BA.2 Variants. N Engl J Med. 2022 Mar 16. PubMed PMID: 35294809. Epub 20220316.

15. Carroll T, Fox D, van Doremalen N, Ball E, Morris MK, Sotomayor-Gonzalez A, et al. The B.1.427/1.429 (epsilon) SARS-CoV-2 variants are more virulent than ancestral B.1 (614G) in Syrian hamsters. PLoS Pathog. 2022 Feb;18(2):e1009914. PubMed PMID: 35143587. Pubmed Central PMCID: PMC8865701. Epub 20220210.

16. Escalera A, Gonzalez-Reiche AS, Aslam S, Mena I, Laporte M, Pearl RL, et al. Mutations in SARS-CoV-2 variants of concern link to increased spike cleavage and virus transmission. Cell Host Microbe. 2022 Mar 9;30(3):373–87 e7. PubMed PMID: 35150638. Pubmed Central PMCID: PMC8776496. Epub 20220121.

17. Halfmann PJ, Iida S, Iwatsuki-Horimoto K, Maemura T, Kiso M, Scheaffer SM, et al. SARS-CoV-2 Omicron virus causes attenuated disease in mice and hamsters. Nature. 2022 Mar;603(7902):687–92. PubMed PMID: 35062015. Epub 20220121.

18. Port JR, Yinda CK, Avanzato VA, Schulz JE, Holbrook MG, van Doremalen N, et al. Increased small particle aerosol transmission of B.1.1.7 compared with SARS-CoV-2 lineage A in vivo. Nat Microbiol. 2022 Feb;7(2):213–23. PubMed PMID: 35017676. Epub 20220111.

19. Saito A, Irie T, Suzuki R, Maemura T, Nasser H, Uriu K, et al. Enhanced fusogenicity and pathogenicity of SARS-CoV-2 Delta P681R mutation. Nature. 2022 Feb;602(7896):300–6. PubMed PMID: 34823256. Pubmed Central PMCID: PMC8828475. Epub 20211125.

20. Ulrich L, Halwe NJ, Taddeo A, Ebert N, Schon J, Devisme C, et al. Enhanced fitness of SARS-CoV-2 variant of concern Alpha but not Beta. Nature. 2022 Feb;602(7896):307–13. PubMed PMID: 34937050. Pubmed Central PMCID: PMC8828469. Epub 20211222.

21. Laskar R, Ali S. Phylo-geo-network and haplogroup analysis of 611 novel coronavirus (SARS-CoV-2) genomes from India. Life Sci Alliance. 2021 May;4(5). PubMed PMID: 33727249. Pubmed Central PMCID: PMC7994317. Epub 2021/03/18.

22. Pattabiraman C, Habib F, P KH, Rasheed R, Prasad P, Reddy V, et al. Genomic epidemiology reveals multiple introductions and spread of SARS-CoV-2 in the Indian state of Karnataka. PLoS One. 2020;15(12):e0243412. PubMed PMID: 33332472. Pubmed Central PMCID: PMC7746284 participate as a volunteer in this study and his employer neither had access to data, nor any say in the design of the study or the decision to publish. This does not alter our adherence to PLOS ONE policies on sharing data and materials. All other authors are employees of state (PD, PK) or central government. The employers had no role in the design of the study or the decision to publish. The authors declare that they do not have any other financial or non-financial relationships that could present a conflict of interest. Epub 2020/12/18.

23. Anantharaj A, Gujjar S, Kumar S, Verma N, Wangchuk J, Khan NA, et al. Kinetics of viral load, immunological mediators and characterization of a SARS-CoV-2 isolate in mild COVID-19 patients during acute phase of infection. medRxiv. 2020:2020.11.05.20226621.

24. Dhar MS, Marwal R, Radhakrishnan V, Ponnusamy K, Jolly B, Bhoyar RC, et al. Genomic characterization and Epidemiology of an emerging SARS-CoV-2 variant in Delhi, India. medRxiv. 2021:2021.06.02.21258076.

25. Zhu N, Wang W, Liu Z, Liang C, Wang W, Ye F, et al. Morphogenesis and cytopathic effect of SARS-CoV-2 infection in human airway epithelial cells. Nat Commun. 2020 Aug 6;11(1):3910. PubMed PMID: 32764693. Pubmed Central PMCID: PMC7413383. Epub 2020/08/09.

26. Liu Y, Liu J, Johnson BA, Xia H, Ku Z, Schindewolf C, et al. Delta spike P681R mutation enhances SARS-CoV-2 fitness over Alpha variant. bioRxiv. 2021 Sep 5. PubMed PMID: 34462752. Pubmed Central PMCID: PMC8404900. Epub 20210905.

27. Mlcochova P, Kemp S, Dhar MS, Papa G, Meng B, Ferreira I, et al. SARS-CoV-2 B.1.617.2 Delta variant replication and immune evasion. Nature. 2021 Sep 6. PubMed PMID: 34488225. Epub 2021/09/07.

28. Li B, Deng A, Li K, Hu Y, Li Z, Xiong Q, et al. Viral infection and transmission in a large, well-traced outbreak caused by the SARS-CoV-2 Delta variant. medRxiv. 2021:2021.07.07.21260122.

29. Suzuki R, Yamasoba D, Kimura I, Wang L, Kishimoto M, Ito J, et al. Attenuated fusogenicity and pathogenicity of SARS-CoV-2 Omicron variant. Nature. 2022 Mar;603(7902):700–5. PubMed PMID: 35104835. Epub 20220201.

30. Bojkova D, Widera M, Ciesek S, Wass MN, Michaelis M, Cinatl J, Jr. Reduced interferon antagonism but similar drug sensitivity in Omicron variant compared to Delta variant of SARS-CoV-2 isolates. Cell Res. 2022 Mar;32(3):319–21. PubMed PMID: 35064226. Pubmed Central PMCID: PMC8781709. Epub 20220121.

31. Zhao H, Lu L, Peng Z, Chen LL, Meng X, Zhang C, et al. SARS-CoV-2 Omicron variant shows less efficient replication and fusion activity when compared with Delta variant in TMPRSS2-expressed cells. Emerg Microbes Infect. 2022 Dec;11(1):277–83. PubMed PMID: 34951565. Pubmed Central PMCID: PMC8774049.

32. Vanderheiden A, Ralfs P, Chirkova T, Upadhyay AA, Zimmerman MG, Bedoya S, et al. Type I and Type III Interferons Restrict SARS-CoV-2 Infection of Human Airway Epithelial Cultures. J Virol. 2020 Sep 15;94(19). PubMed PMID: 32699094. Pubmed Central PMCID: PMC7495371. Epub 2020/07/24.

33. Schroeder S, Pott F, Niemeyer D, Veith T, Richter A, Muth D, et al. Interferon antagonism by SARS-CoV-2: a functional study using reverse genetics. Lancet Microbe. 2021 May;2(5):e210–e8. PubMed PMID: 33969329. Pubmed Central PMCID: PMC8096348. Epub 20210304.

34. Anantharaj A, Gujjar S, Verma N, Khan NA, Shaman H, Sharanabasava P, et al. Resolution of viral load in mild COVID-19 patients is associated with both innate and adaptive immune responses. Journal of Clinical Virology. 2022 2022/01/01/;146:105060.

35. Bojkova D, Rothenburger T, Ciesek S, Wass MN, Michaelis M, Cinatl J. SARS-CoV-2 Omicron variant virus isolates are highly sensitive to interferon treatment. bioRxiv. 2022:2022.01.20.477067.

36. Desingu PA, Nagarajan K, Dhama K. Emergence of Omicron third lineage BA.3 and its importance. Journal of Medical Virology. 2022;94(5):1808–10.

37. Medigeshi G, Batra G, Murugesan DR, Thiruvengadam R, Chattopadhyay S, Das B, et al. Sub-optimal Neutralisation of Omicron (B.1.1.529) Variant by Antibodies induced by Vaccine alone or SARS-CoV-2 Infection plus Vaccine (Hybrid Immunity) post 6-months. medRxiv. 2022:2022.01.04.22268747.

38. Lyngse FP, Kirkeby CT, Denwood M, Christiansen LE, Mølbak K, Møller CH, et al. Transmission of SARS-CoV-2 Omicron VOC subvariants BA.1 and BA.2: Evidence from Danish Households. medRxiv. 2022:2022.01.28.22270044.

39. Yamasoba D, Kimura I, Nasser H, Morioka Y, Nao N, Ito J, et al. Virological characteristics of SARS-CoV-2 BA.2 variant. bioRxiv. 2022:2022.02.14.480335.

40. Arora P, Zhang L, Rocha C, Sidarovich A, Kempf A, Schulz S, et al. Comparable neutralisation evasion of SARS-CoV-2 omicron subvariants BA.1, BA.2, and BA.3. The Lancet Infectious Diseases.

41. Chaturvedi UC, Shrivastava R. Interaction of viral proteins with metal ions: role in maintaining the structure and functions of viruses. FEMS Immunol Med Microbiol. 2005 Feb 1;43(2):105–14. PubMed PMID: 15681139. Pubmed Central PMCID: PMC7110337.

42. e Velthuis AJ, van den Worm SH, Sims AC, Baric RS, Snijder EJ, van Hemert MJ. Zn(2+) inhibits coronavirus and arterivirus RNA polymerase activity in vitro and zinc ionophores block the replication of these viruses in cell culture. PLoS Pathog. 2010 Nov 4;6(11):e1001176. PubMed PMID: 21079686. Pubmed Central PMCID: PMC2973827. Epub 2010/11/17.

43. Hillen HS, Kokic G, Farnung L, Dienemann C, Tegunov D, Cramer P. Structure of replicating SARS-CoV-2 polymerase. Nature. 2020 Aug;584(7819):154–6. PubMed PMID: 32438371. Epub 20200521.

44. Shimizu H, Saito A, Mikuni J, Nakayama EE, Koyama H, Honma T, et al. Discovery of a small molecule inhibitor targeting dengue virus NS5 RNA-dependent RNA polymerase. PLOS Neglected Tropical Diseases. 2019;13(11):e0007894.

45. Eydoux C, Fattorini V, Shannon A, Le TT, Didier B, Canard B, et al. A fluorescence-based high throughput-screening assay for the SARS-CoV RNA synthesis complex. J Virol Methods. 2021 Feb;288:114013. PubMed PMID: 33166547. Pubmed Central PMCID: PMC7833800. Epub 20201106.

46. Nealon J, Cowling BJ. Omicron severity: milder but not mild. The Lancet. 2022;399(10323):412–3.

47. Dan JM, Mateus J, Kato Y, Hastie KM, Yu ED, Faliti CE, et al. Immunological memory to SARS-CoV-2 assessed for up to 8 months after infection. Science. 2021 Feb 5;371(6529). PubMed PMID: 33408181. Pubmed Central PMCID: PMC7919858. Epub 20210106.

48. Chen J, Wei GW. Omicron BA.2 (B.1.1.529.2): high potential to becoming the next dominating variant. ArXiv. 2022 Feb 10. PubMed PMID: 35169598. Pubmed Central PMCID: PMC8845508. Epub 20220210.

49. Gao Y, Cai C, Grifoni A, Muller TR, Niessl J, Olofsson A, et al. Ancestral SARS-CoV-2-specific T cells cross-recognize the Omicron variant. Nat Med. 2022 Jan 14. PubMed PMID: 35042228. Epub 20220114.

50. GeurtsvanKessel CH, Geers D, Schmitz KS, Mykytyn AZ, Lamers MM, Bogers S, et al. Divergent SARS CoV-2 Omicron-reactive T-and B cell responses in COVID-19 vaccine recipients. Sci Immunol. 2022 Feb 3:eabo2202. PubMed PMID: 35113647. Epub 20220203.

51. Broggi A, Ghosh S, Sposito B, Spreafico R, Balzarini F, Lo Cascio A, et al. Type III interferons disrupt the lung epithelial barrier upon viral recognition. Science. 2020 Aug 7;369(6504):706–12. PubMed PMID: 32527925. Pubmed Central PMCID: PMC7292499. Epub 2020/06/13.

52. Major J, Crotta S, Llorian M, McCabe TM, Gad HH, Priestnall SL, et al. Type I and III interferons disrupt lung epithelial repair during recovery from viral infection. Science. 2020 Aug 7;369(6504):712–7. PubMed PMID: 32527928. Pubmed Central PMCID: PMC7292500. Epub 2020/06/13.

53. Hui KPY, Ho JCW, Cheung MC, Ng KC, Ching RHH, Lai KL, et al. SARS-CoV-2 Omicron variant replication in human bronchus and lung ex vivo. Nature. 2022 Mar;603(7902):715–20. PubMed PMID: 35104836. Epub 20220201.

54. McMahan K, Giffin V, Tostanoski LH, Chung B, Siamatu M, Suthar MS, et al. Reduced pathogenicity of the SARS-CoV-2 omicron variant in hamsters. Med. 2022 2022/04/08/;3(4):262–8.e4.

55. Wang H, Li Z, Niu J, Xu Y, Ma L, Lu A, et al. Antiviral effects of ferric ammonium citrate. Cell Discov. 2018;4:14. PubMed PMID: 29619244. Pubmed Central PMCID: PMC5871618. Epub 20180327.

56. Maio N, Lafont BAP, Sil D, Li Y, Bollinger JM, Jr., Krebs C, et al. Fe-S cofactors in the SARS-CoV-2 RNA-dependent RNA polymerase are potential antiviral targets. Science. 2021 Jul 9;373(6551):236–41. PubMed PMID: 34083449. Pubmed Central PMCID: PMC8892629. Epub 20210603.

57. Bastin A, Shiri H, Zanganeh S, Fooladi S, Momeni Moghaddam MA, Mehrabani M, et al. Iron Chelator or Iron Supplement Consumption in COVID-19? The Role of Iron with Severity Infection. Biol Trace Elem Res. 2021 Nov 25. PubMed PMID: 34825316. Pubmed Central PMCID: PMC8614629. Epub 20211125.

58. Habib HM, Ibrahim S, Zaim A, Ibrahim WH. The role of iron in the pathogenesis of COVID-19 and possible treatment with lactoferrin and other iron chelators. Biomed Pharmacother. 2021 Apr;136:111228. PubMed PMID: 33454595. Pubmed Central PMCID: PMC7836924. Epub 20210113.

59. Bryant C, Wilks SA, Keevil CW. Rapid inactivation of SARS-CoV-2 on copper touch surfaces determined using a cell culture infectivity assay. bioRxiv. 2021:2021.01.02.424974.

60. Govind V, Bharadwaj S, Sai Ganesh MR, Vishnu J, Shankar KV, Shankar B, et al. Antiviral properties of copper and its alloys to inactivate covid-19 virus: a review. BioMetals. 2021 2021/12/01;34(6):1217–35.

61. Raha S, Mallick R, Basak S, Duttaroy AK. Is copper beneficial for COVID-19 patients? Medical Hypotheses. 2020 2020/09/01/;142:109814.

62. Sagripanti JL, Routson LB, Lytle CD. Virus inactivation by copper or iron ions alone and in the presence of peroxide. Appl Environ Microbiol. 1993 Dec;59(12):4374–6. PubMed PMID: 8285724. Pubmed Central PMCID: PMC195916.

63. Viswanathan T, Misra A, Chan SH, Qi S, Dai N, Arya S, et al. A metal ion orients SARS-CoV-2 mRNA to ensure accurate 2’-O methylation of its first nucleotide. Nat Commun. 2021 Jun 2;12(1):3287. PubMed PMID: 34078893. Pubmed Central PMCID: PMC8172916. Epub 20210602.

64. Bouvet M, Lugari A, Posthuma CC, Zevenhoven JC, Bernard S, Betzi S, et al. Coronavirus Nsp10, a critical co-factor for activation of multiple replicative enzymes. J Biol Chem. 2014 Sep 12;289(37):25783–96. PubMed PMID: 25074927. Pubmed Central PMCID: PMC4162180. Epub 20140729.

65. Chen T, Fei CY, Chen YP, Sargsyan K, Chang CP, Yuan HS, et al. Synergistic Inhibition of SARS-CoV-2 Replication Using Disulfiram/Ebselen and Remdesivir. ACS Pharmacol Transl Sci. 2021 Apr 9;4(2):898–907. PubMed PMID: 33855277. Pubmed Central PMCID: PMC8009100. Epub 20210326.

66. Anantharaj A, Das SJ, Sharanabasava P, Lodha R, Kabra SK, Sharma TK, et al. Visual Detection of SARS-CoV-2 RNA by Conventional PCR-Induced Generation of DNAzyme Sensor. Front Mol Biosci. 2020 2020-December-23;7(444):586254. PubMed PMID: 33425988. Pubmed Central PMCID: PMC7793695. Epub 2021/01/12. English.

67. Das S, Singh J, Shaman H, Singh B, Anantharaj A, Sharanabasava P, et al. Antibody response after a single dose of BBV152 vaccine negatively correlates with pre-existing antibodies and induces a significant but low levels of neutralizing antibodies to Omicron variant. medRxiv. 2022:2022.02.07.22270612.

