## Supplementary material for "BA.1 and BA.2 sub-lineages of Omicron variant have comparable replication kinetics and susceptibility to neutralization by antibodies": Combined Supplementary data

### SUPPLEMENTARY METHODS

#### Whole genome sequencing of SARS-CoV-2 using Nanopore platform

Genome sequencing was performed according to Oxford Nanopore Technology (ONT) library preparation protocol- PCR tiling of SARS-CoV-2 virus with Rapid barcoding (Version: PCTR_9125_v110_revB_24Mar2021), commonly known as the midnight protocol. 50 ng of the extracted RNA from nasopharyngeal samples was reverse transcribed into cDNA using LunaScript RT SuperMix, comprising of random hexamer and oligo-dT primers, dNTPs, Murine RNase Inhibitor, and Reverse Transcriptase. The cDNA-RNA hybrid was used to amplify SARS-CoV-2 genome with rapid barcoding primers (IDT Product number: 10007184) and Q5 High-Fidelity 2X master mix (New England Biolabs, Cat. No. M0494S). The primer pools consisted of 30 and 28 primers each, that were designed specifically to produce 1200 bp amplicons with an approximate overlap of 20 bp. Targeted amplification, using multiplex PCR, was performed for each SARS-CoV-2 positive sample, separately for both the primer pools. The amplified products for both the primer pools were then combined and ligated to unique barcode sequences (SQK-RBK110.96). The barcode ligation step required a Transposase which simultaneously fragmented 1200 bp long amplicons and added barcode sequences to the cleaved ends. The barcoded library was pooled and then purified using SPRI beads. Tagging of unique barcode to each sample enabled pooling of multiple samples for sequencing together in the same library. The pooled barcoded library was quantified using Qubit dsDNA HS Assay kit. Finally, 800 ng of the library was then ligated with adapter protein and loaded on MinION Mk1B or MinION Mk1C platform.

#### Nanopore Analysis Method

The ARTIC end-to-end pipeline was used for the analysis of ONT MinION raw fast5 files up to variant calling. Raw fast5 files of samples was base called and demultiplexed using barcode kits information SQK-RBK110-96 with Guppy base caller that uses the base calling algorithms of Oxford Nanopore Technologies (Nanopore Community) with phred quality cut-off score >7 on GPU-linux accelerated computing machine. Reads having Phred quality score less than 7 were discarded to filter the low-quality reads. The resultant demultiplexed fastq were normalized by read length using the 1200 bp amplicon sets for further downstream analysis and aligned to the SARS-CoV-2 reference (MN908947.3) using the aligner Minimap2. Nanopolish were used to index raw fast5 files for variant calling from the minimap output files. To create consensus fasta, bcftools was used with normalized minimap2 output. Further, the assembled SARS-CoV-2 genomes were assigned lineages using the package Phylogenetic Assignment of Named Global Outbreak LINeages (PANGOLIN).

**Cell viability assay**

Cell viability assay was performed using CellTiter-Glo Assay kit as per the manufacturer's instructions. This method determines the number of viable cells in culture based on quantification of the ATP present, in metabolically active cells. Briefly, Calu-3 cells were seeded at a density of 50,000 in 48-well plate and grown for 48 h. Cells were treated with indicated metal ions at a concentration of 50 μM in a total volume of 300 μl serum-free growth medium. After 24 h post-treatment, the assay was performed. 200 μl of medium was discarded from each well.100 μl of CellTiter-Glo reagent was added to the 100 μl of medium containing cells in each well. The contents were mixed for 2 min on rocker for cell lysis. The plate was incubated at room temperature for 10 min. The homogeneous CellTiter-Glo reagent results in cell lysis and generates a luminescent signal proportional to the amount of ATP present. 100 µl of content from each well was transferred to opaque-walled multiwell plates. The luminescence was measured by microplate reader (Synergy HT - BioTek).

### KEY RESOURCES TABLE

| REAGENTS OR RESOURCES | SOURCE | IDENTIFIER |
|---|---|---|
| **Antibodies** | | |
| SARS-CoV-2 Nucleocapsid | GenScript | Catalog no. A02048 |
| SARS-CoV-2 Spike RBD | Sino Biological | Catalog no. 40592-T62 |
| β-catenin | BD Biosciences | Catalog no. 610154 |
| Occludin | Invitrogen | Catalog no. 71-1500 |
| GAPDH | Cell Signaling Technology | Catalog no. 2118S |
| Alexa fluor 488 goat anti-human IgG | Invitrogen | Catalog no. A-11013 |
| Alexa fluor 488 goat anti-mouse IgG2b | Invitrogen | Catalog no. A-21141 |
| Alexa fluor 633 goat anti-mouse IgG1 | Invitrogen | Catalog no. A-21126 |
| Alexa fluor 568 donkey anti-rabbit IgG | Invitrogen | Catalog no. A-10042 |
| Goat Anti-Rabbit IgG (H+L) Horseradish Peroxidase conjugate | Invitrogen | Catalog no. G-21234 |
| Goat anti-Mouse IgG (H+L) Cross-Adsorbed Secondary Antibody, HRP | Invitrogen | Catalog no. A16072 |
| HRP-Goat Anti- Human IgG | Jackson ImmunoResearch | Catalog no. 109-035-170 |
| **Virus strains** | | |
| BL-THSTI 2010D (B.6) | GenBank accession: MW422884.1 | |
| Kappa (B.1.617.1) | GenBank accession: MZ356902.1 | |
| Delta (B.1.617.2) | GenBank accession: MZ356566.1 | |
| Omicron BA.1 | GISAID-accession no EPI_ISL_6716890 | |
| Omicron BA.2 | GISAID-accession no EPI_ISL_87638432 | |
| **Media and chemicals** | | |
| Dulbecco's Modified Eagle Medium (DMEM), High glucose | HiMedia | Catalog no. AL007A |
| Minimum Essential Media (MEM) | Thermo Fisher/Gibco | Catalog no. 11090073 |
| Penicillin-Streptomycin-Glutamine (PSG) 100X | Thermo Fisher | Catalog no. 10378016 |
| Antibiotic solution 100X liquid | HiMedia | Catalog no. A001 |
| Non-Essential Amino Acids Solution (100X) | Thermo Fisher | Catalog no. 11140-050 |
| Fetal Bovine Serum (FBS) | Thermo Fisher | Catalog no. 16140-071 |

| | | |
|---|---|---|
| Trypsin-EDTA (0.25%), phenol red | Thermo Fisher | Catalog no. 25200072 |
| 6.5 mm Transwell® with 3.0 µm Pore Polycarbonate Membrane Insert | Corning | Catalog no. 3415 |
| PrimeScript RT reagent kit with gDNA eraser kit | TaKaRa | Catalog no. RR047A |
| PowerUp™ SYBR™ Green Master Mix | Thermo Fisher | Catalog no. A25742 |
| ProLong™ Gold Antifade Mountant | Thermo Fisher | Catalog no. P36934 |
| 1x Dulbecco's Phosphate Buffered Saline (DPBS) | Thermo Fisher/Gibco | Catalog no. 21600010 |
| Carboxymethylcellulose (CMC) medium viscosity | Sigma | Catalog no. C4888 |
| Methanol | Merck | Catalog no. SB0SF70138 |
| Paraformaldehyde | Sigma-Aldrich | Catalog no. P6148 |
| HEPES | HiMedia | Catalog no. MB016 |
| 2019-nCoV CDC Probe and Primer kit for SARS-CoV-2 | Biosearch Technologies | Catalog no. KIT-nCoV-PP1-1000 |
| TaqMan™ RNase P Assay, VIC™ dye/QSY™ probe | Thermo Fisher | Catalog no: A30064 |
| Protease inhibitor cocktail (PIC) | Merck | Catalog no. 11836170001 |
| Phenylmethylsulfonyl fluoride (PMSF) | Sigma-Aldrich | Catalog no. P7626 |
| PVDF membrane | GE Healthcare Life Sciences | Catalog no. 10600023 |
| RNA Isolation kit | Macherey-Nagel | Catalog no. 740955.250 |
| CellTiter-Glo assay kit | Promega | Catalog no. 7571 |
| Quant-iT™ PicoGreen™ dsDNA Assay Kit | Thermo-Fisher | Catalog no. 7589 |
| **Cell lines** | | |
| Vero E6 | ECACC | Catalog no. 85020206 |
| Calu-3 cells | ATCC | Catalog no. ATCC-HTB-55 |
| **Primers** | | |
| **Gene** | **Forward primer (5’-3’)** | **Reverse primer (5’-3’)** |
| *GAPDH* | CCACTCCTCCACCTTTGAC | ACCCTGTTGCTGTAGCCA |
| *IFN-β* | AAACTCATGAGCAGTCTGCA | AGGAGATCTTCAGTTTCGGAGG |
| *IFN-λ1* | GGTGACTTTGGTGCTAGGCT | TGAGTGACTCTTCCAAGGCG |
| *ISG15* | AGATCACCCAGAAGATCG | TGTTATTCCTCACCAGGATG |
| *OAS1* | TTCAGCGAGCTGCAGAGAAA | AAGAGCATAGAGAGGGGGCA |
| **Software and Algorithms** | | |
| Prism (9.1.0) | GraphPad | |
| cellSens | Olympus | |
| AID EliSpot 8.0 iSpot software | AID | |
| Gen5 | BioTek | |
| SoftMax pro GXP 7.1 | Molecular Devices | |

### SUPPLEMENTARY FIGURE LEGENDS

**Supplementary Figure S1. COVID-19 diagnostic testing.** (A) NP/OP samples were tested by RT-PCR to diagnose COVID-19 infection and % positivity (i) and number of positive cases relative to the total number of samples tested (ii) from April 2020 to July 2021 is shown. (B) Threshold cycle (Ct) values of RNA samples tested for COVID-19 during the same period is shown and the values are segregated into four categories to indicate very high (< 20 Ct), high (20-25 Ct), moderate (25-30 Ct) and low (>30 Ct) viral burden in the first sample collected for diagnosis.

**Supplementary Figure S2: Distribution of SARS-CoV-2 lineages in National Capital Region of India between the months of September 2021 to January 2022.** Whole genome sequencing of COVID-19 positive diagnostic samples for the indicated period. B.1.617.2 (Delta); AY.* (Delta plus); Omicron lineages (B.1.529, BA.1 and BA.2).

**Supplementary Figure S3: Growth characteristics of Omicron variant and establishment of FRNT assay:** (A) Calu-3 or Vero E6 cells were incubated with 10-fold serial dilution of Omicron (BA.1) variant to determine virus titers by plaque assay. Plates were fixed at 24 and 48 h pi and stained with crystal violet. (B) Vero E6 cells were infected with a pre-determined dilution of Omicron variant for focus-forming unit assay using anti-spike and anti-nucleocapsid antibodies followed by HRP-conjugated secondary antibody. Foci were developed using TrueBlue substrate. (C) Vero E6 cells were infected with a pre-determined dilution of Omicron variant for focus-forming unit assay using anti-spike and anti-nucleocapsid antibodies followed by Alexa488-conjugated secondary antibody. Foci were visualized under fluorescence channel in the reader. (D) FRNT assay using fluorescence method to determine the neutralization titers of antibodies against the Delta and Omicron variants. 50% neutralization titer of antibodies ($NT_{50}$) is given from at least five experimental replicates (Mean ± SD).

**Supplementary Figure S4: Effect of divalent cations on cell viability.** Calu-3 cells were treated with indicated salts at a concentration of 50 µM for 24 h. Cell viability assay was performed using CellTiter-Glo luminescent cell viability assay. Data from two experiments are presented as Mean + SD.

### Supplementary Figure S1

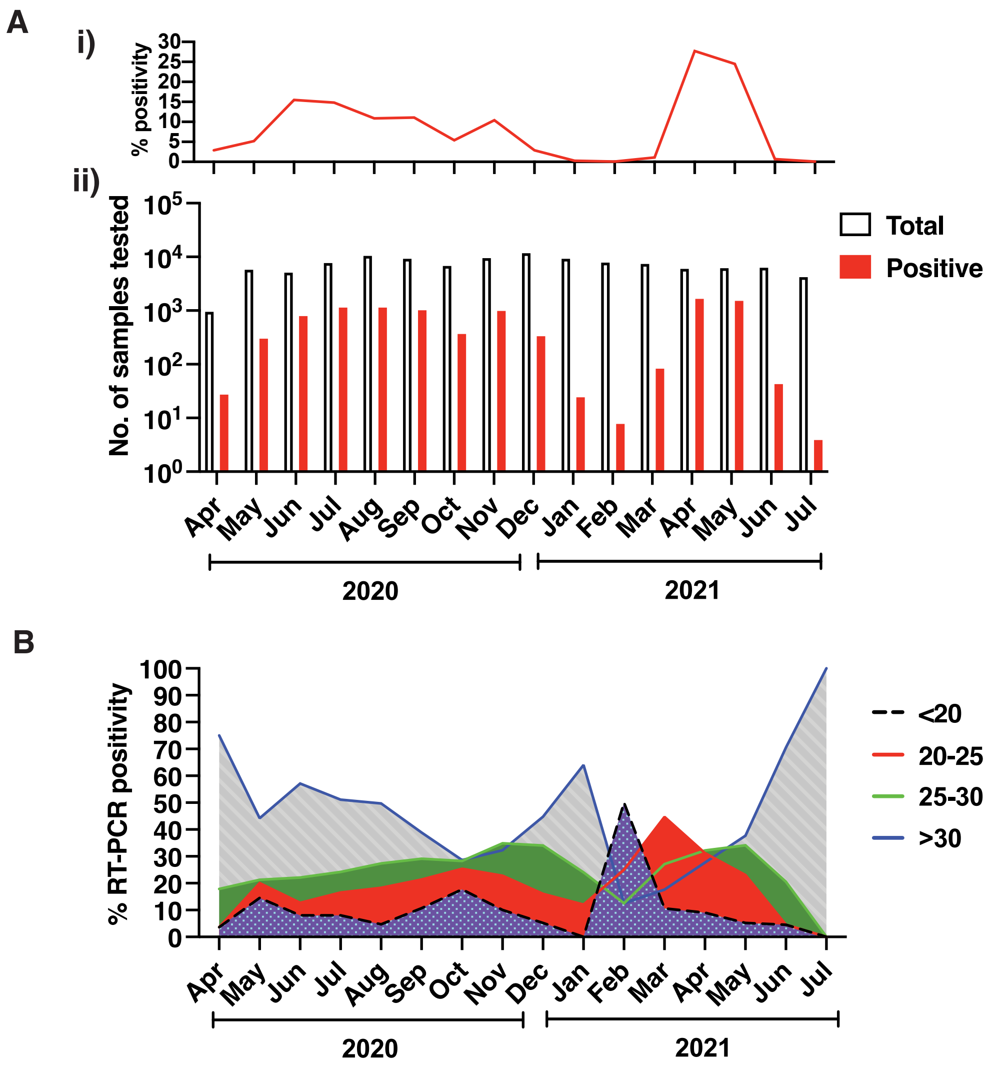

**Supplementary Fgure S1: COVID-19 diagnostic testing.**

### Supplementary Figure S2

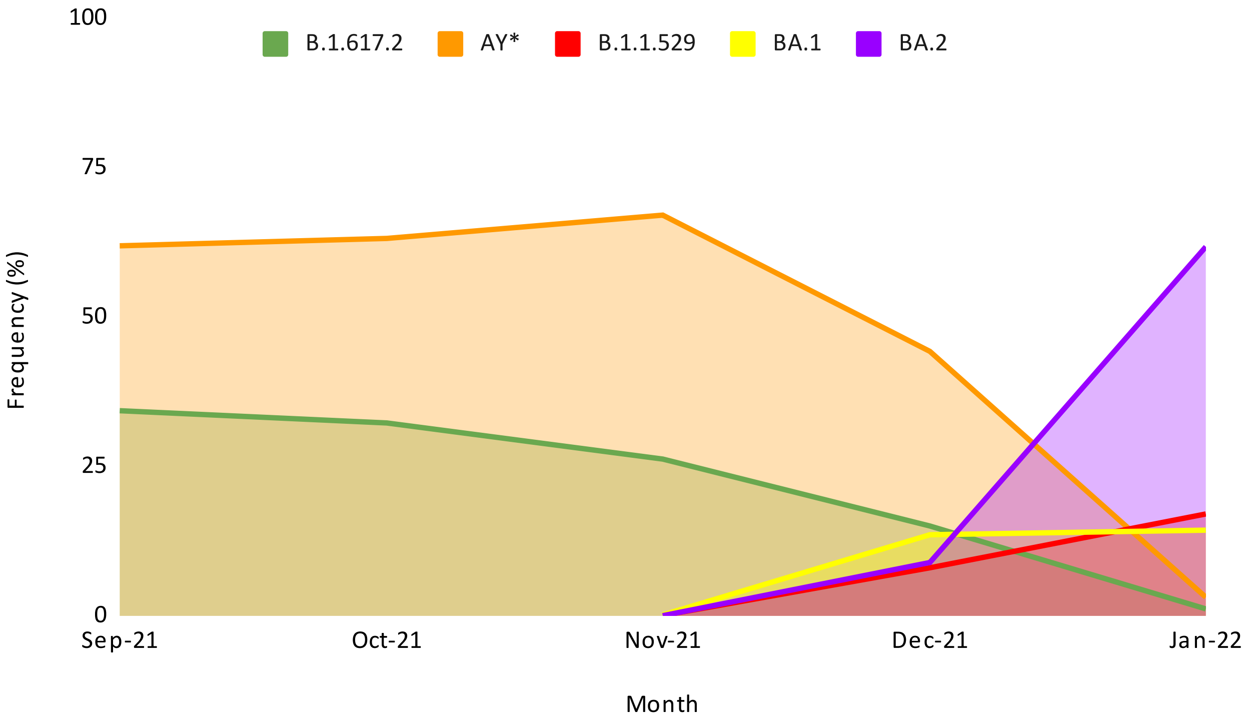

**Supplementary Fgure S2: Distribution of SARS-CoV-2 lineages in National Capital Region of India between the months of September 2021 to January 2022.**

### Supplementary Figure S3

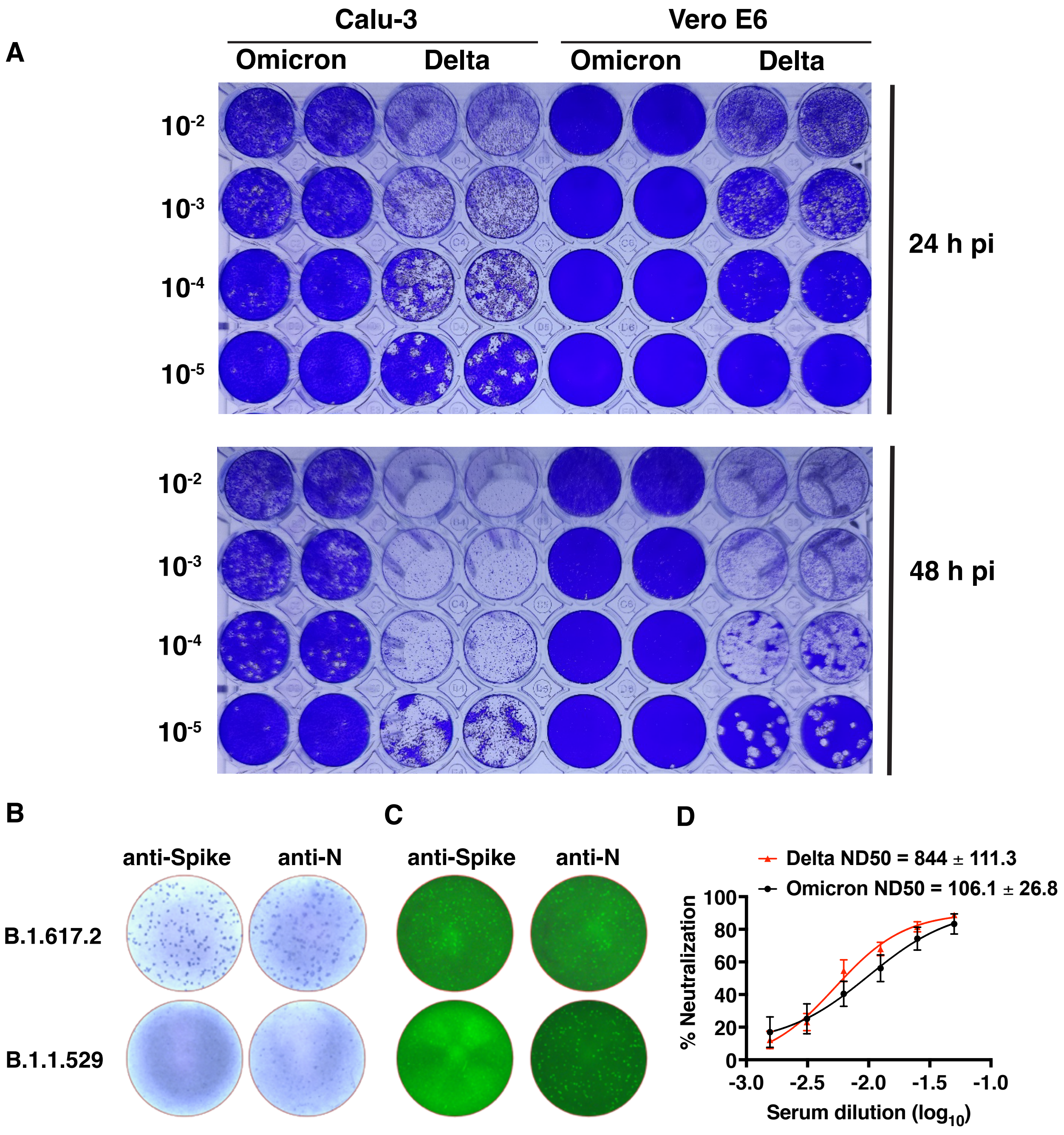

**Supplementary Fgure S3: Growth characteristics of Omicron variant and establishment of FRNT assay**

### Supplementary Figure S4

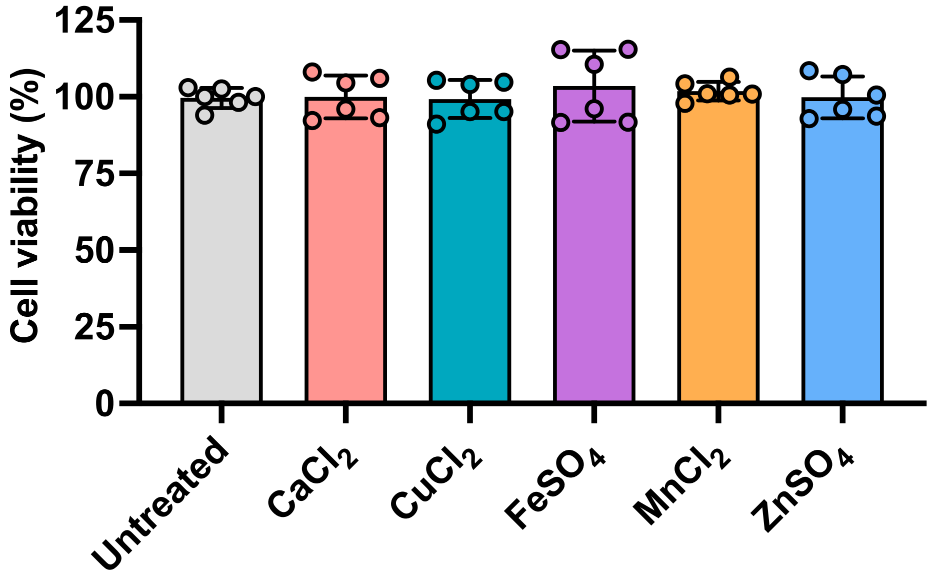

**Supplementary Fgure S4: Viability of cells treated with salts of divalent cations.**
